# CagriSema 2.4/2.4 mg versus CagriSema 1.0/1.0 mg in adults with type 2 diabetes: a dose-comparison systematic review and meta-analysis of randomized controlled trials

**DOI:** 10.64898/2026.10.30.26366916

**Authors:** Bernardo Madeira Diefenthaeler, Livia Eichenberg do Canto, Gabriela Menegaz Pretto, Henrique Corleto Malta, Guilherme Lopes Noll, Luiza de Oliveira Magalhães, Gabriela Gonzalez dos Santos, Fernando Gerchman

## Abstract

**Background:** Once-weekly cagrilintide–semaglutide (CagriSema) has been evaluated at two maintenance doses, 2.4/2.4 mg and 1.0/1.0 mg, in type 2 diabetes (T2DM), but they have not been directly compared. We compared their efficacy and safety.

**Methods:** We searched PubMed, Embase, and Cochrane CENTRAL for randomized controlled trials (RCTs) comparing both doses. Contrasts derived from arm-level estimates were pooled using random-effects models. Primary outcomes were interpreted against prespecified minimally important differences (MIDs) of 3 percentage points (pp) for body weight and 0.3 pp (3.3 mmol/mol) for glycated hemoglobin (HbA1c).

**Results:** Three trials (1,506 participants) were included. CagriSema 2.4/2.4 mg produced a greater reduction in body weight (mean difference [MD] −2.07 pp; 95% confidence interval [CI] −2.91 to −1.24), with the entire CI below the MID, and higher proportions achieving reductions of ≥10% (64.9% vs. 55.1%) and ≥15% (41.9% vs. 30.9%). The difference in HbA1c was not statistically significant (MD −0.16 pp [−1.7 mmol/mol]; 95% CI −0.43 to 0.10 [−4.7 to 1.1 mmol/mol]). The higher dose increased gastrointestinal disorders (64.8% vs. 53.2%; risk ratio 1.22; 95% CI 1.12 to 1.32), without differences in serious adverse events or adverse events leading to treatment discontinuation. Certainty of evidence was high for body weight and moderate for HbA1c.

**Conclusions:** CagriSema 2.4/2.4 mg provided additional weight loss smaller than the MID, no clear HbA1c benefit, and more gastrointestinal adverse events than 1.0/1.0 mg, supporting individualized dose selection.

**Registration:** CRD420261545958 (registered 10/21/2026).

## Introduction

Overweight and obesity are major public health challenges and are strongly associated with insulin resistance, metabolic syndrome, and type 2 diabetes mellitus (T2DM) [1, 2]. According to the World Health Organization, 2.6 billion adults were living with overweight in 2024, including 940 million with obesity, and approximately 830 million people live with diabetes [3, 4]. Consequently, novel pharmacological therapies that promote both weight loss and glycemic control have been developed [5, 6].

CagriSema is a fixed-dose combination of cagrilintide, a long-acting amylin analogue that delays gastric emptying and increases satiety, and semaglutide, a glucagon-like peptide-1 (GLP-1) receptor agonist that also stimulates insulin secretion and suppresses glucagon release [7–9]. Their complementary mechanisms of action have produced greater metabolic effects than either agent alone in people with overweight or obesity [10–12]. Following a phase 1b dose-finding trial [9], the combination of cagrilintide 2.4 mg and semaglutide 2.4 mg became the most extensively studied regimen in subsequent trials, showing a favorable response across diverse patient populations [12–14].

Despite these benefits, the dose-dependent benefit–risk profile of CagriSema remains insufficiently characterized. Semaglutide monotherapy target doses have evolved according to clinical indication: for T2DM, the United States Food and Drug Administration (FDA) initially approved a maximum maintenance dose of 1.0 mg in 2017 [15], later increased to 2.0 mg in 2022 following the SUSTAIN FORTE trial [16, 17]. For chronic weight management, doses of 2.4 mg and, more recently, 7.2 mg were approved [18, 19]. In contrast, CagriSema has not yet been approved, and no standard dose has been established for people with T2DM. Although recent randomized controlled trials have evaluated both the 1.0/1.0 mg and 2.4/2.4 mg doses [20–22], existing meta-analyses have either pooled data for a single dose or compared each dose separately with other comparators [10, 23, 24]. A head-to-head synthesis of these doses is therefore warranted.

This systematic review and meta-analysis aims to evaluate the comparative efficacy and safety of CagriSema doses of 1.0/1.0 mg versus 2.4/2.4 mg in adults with T2DM. We aim to evaluate their relative effects on weight loss and glycemic control, as well as adverse effects and other metabolic outcomes.

## Methods

This systematic review and meta-analysis was conducted in accordance with the Cochrane Handbook for Systematic Reviews of Interventions and reported following the Preferred Reporting Items for Systematic Reviews and Meta-Analyses (PRISMA) 2020 guidelines [25, 26] (checklist in Supplementary File 1). The protocol was prospectively registered in the International Prospective Register of Systematic Reviews (PROSPERO; CRD420261545958).

### Eligibility criteria

Studies were considered eligible if they met the following criteria: (1) randomized controlled trials (RCTs); (2) enrollment of adults (≥18 years) with T2DM; and (3) comparison of once-weekly subcutaneous cagrilintide 2.4 mg plus semaglutide 2.4 mg (CagriSema 2.4/2.4 mg) with cagrilintide 1.0 mg plus semaglutide 1.0 mg (CagriSema 1.0/1.0 mg), regardless of the dose-escalation regimen.

Multi-arm trials were eligible when at least one intervention arm and one comparator arm met the prespecified criteria; only those arms were included in the meta-analysis. Studies of combinations in which only one component varied in dose were excluded, as were protocols, conference abstracts, and registry records without sufficient data for extraction.

### Search strategy and data extraction

A systematic search of PubMed, Embase, and the Cochrane Central Register of Controlled Trials (CENTRAL) was conducted from the inception of each database to October 19, 2026, without language or publication date restrictions. The search strategy combined terms related to cagrilintide, semaglutide, and the CagriSema co-formulation with a filter for randomized trials; the complete database-specific strategies are provided in Supplementary File 2 (Table S1). The reference lists of the included studies were also screened manually. After removal of duplicates, titles and abstracts, followed by full texts, were screened independently by two reviewers (GLN and GGS), with disagreements resolved by a third reviewer (BMD).

Data were extracted by two authors (LOM and GGS for study characteristics, and BMD and GLN for outcome data) using a standardized spreadsheet. Discrepancies were resolved by consensus. Extracted characteristics included background glucose-lowering therapy, treatment duration, sample size per arm, sex, age, diabetes duration, baseline HbA1c, body weight, and body mass index. For each outcome, depending on its type, we extracted arm-level means and standard errors (SEs), estimated treatment effects with 95% confidence intervals (CIs), or the number of participants with events. Data corresponded to the end of the randomized treatment period and to the efficacy (trial product) estimand, the primary estimand of all included trials, as prespecified in the protocol.

### Outcomes

The primary outcomes were the percentage change in body weight (percentage points [pp]) and the change in glycated hemoglobin (HbA1c, pp). HbA1c values were additionally expressed in mmol/mol using the standard conversion relationship [27]. The main secondary efficacy outcomes were fasting plasma glucose (mmol/L), waist circumference (cm), systolic blood pressure (mmHg), total cholesterol, low-density lipoprotein cholesterol (LDL-C), and the proportions of participants achieving body-weight reductions of ≥10% and ≥15% from baseline. The main safety outcomes were any adverse event, serious adverse events, gastrointestinal disorders, and adverse events leading to permanent treatment discontinuation. As prespecified in the protocol, additional efficacy, tolerability, and safety outcomes reported in a comparable manner were extracted and meta-analyzed whenever reported by at least three studies; the complete list is presented in the Results. Adverse events were analyzed as the number of participants with at least one event, according to each study’s definitions, and were synthesized even when one of the trials reported no events.

### Statistical analysis

Meta-analyses were conducted using a random-effects model, given the anticipated clinical heterogeneity across the included trials, with the between-study variance (τ²) estimated using the restricted maximum-likelihood (REML) method. When τ² > 0, the CI of the pooled estimate was calculated with the Hartung–Knapp–Sidik–Jonkman (HKSJ) adjustment; when τ² was estimated at zero (including estimates <0.0001), a Wald-type CI was used [25]. All effects were expressed for CagriSema 2.4/2.4 mg relative to CagriSema 1.0/1.0 mg (reference).

Because no trial reported a direct comparison between the two doses, each study-level contrast was constructed from arm-level estimates, an approach also adopted in previous meta-analyses on this topic [10, 28]. For continuous outcomes, the mean difference (MD) was calculated as the difference between the model-adjusted means of the two arms and pooled using the inverse-variance method. The SE of each arm was converted to an effective standard deviation (SD = SE × √N, where N is the number of randomized participants) [25]. As the covariance between the two arm-level means was not reported, the SE of the MD was calculated assuming independence (SE(MD) = √[SE(2.4)² + SE(1.0)²]). When a trial reported only the arm-level mean together with the 95% CI of the difference versus the corresponding semaglutide dose [22], the arm-level SE was derived from that CI under the same independence assumption and assuming equal SDs in both arms (SD = SE of the difference/√[1/N_CagriSema + 1/N_semaglutide]; SE = SD/√N_CagriSema) [25]. Detailed derivations are provided in Supplementary File 2 (Supplementary Methods S1).

For lipid parameters and high-sensitivity C-reactive protein, the trials reported the geometric mean ratio of the end-of-treatment to the baseline value. Because this measure is multiplicative and its sampling distribution is approximately normal only on the logarithmic scale, the analyses were performed on the natural log scale (ln[ratio₂.₄] − ln[ratio₁.₀]). The SE of each arm was obtained from the logarithm of the multiplicative SE or, when this was unavailable or reported with insufficient precision, from the 95% CI of the estimated treatment ratio versus semaglutide, as described above. Pooled estimates were obtained using the generic inverse-variance method and expressed as the ratio of geometric means (RoM).

For binary outcomes, risk ratios (RRs) with 95% CIs were calculated using the Mantel–Haenszel method; in the random-effects model, study-specific estimates were combined with inverse-variance weights, and a continuity correction of 0.5 was applied to studies with zero events in one arm. Studies with zero events in both arms did not contribute to the pooled estimate but were retained in the forest plots. For events with an overall incidence <1%, the Peto odds ratio was used under a common-effect model. For body-weight and HbA1c targets, reported as model-estimated proportions, the number of events was approximated as the product of the proportion and the number of randomized participants in each arm.

Statistical heterogeneity was assessed using Cochran’s Q test, τ², and the I² statistic, the latter calculated from the REML estimate of τ². As an additional interpretive step, the pooled differences in the primary outcomes were compared with prespecified minimally important differences (MIDs): 3 pp for body-weight change, corresponding to the additional reduction observed with semaglutide 2.4 mg versus 1.0 mg in adults with T2DM [29], and 0.3 pp (3.3 mmol/mol) for HbA1c, the non-inferiority margin generally considered acceptable by the agencies’ guidelines [30, 31]. Differences with a point estimate equal to or greater than the MID were considered clinically important in favor of CagriSema 2.4/2.4 mg; when the entire 95% CI lay below the MID, the 1.0/1.0 mg dose was considered not meaningfully inferior; all other results were considered inconclusive. This interpretation does not constitute formal non-inferiority testing.

All statistical analyses were conducted in R, version 4.5.2, using the meta package (version 8.3-0) [32, 33].

### Sensitivity analyses

For outcomes with τ² > 0, a Wald-type CI was additionally calculated as a sensitivity analysis. Because the HKSJ adjustment may yield excessively wide intervals when only three studies are available, the Wald-type interval might be preferred when all studies indicate an effect in the same direction [25]. For the primary outcomes, the analyses were also repeated using data from each trial’s treatment-policy estimand.

### Risk-of-bias assessment

The risk of bias of each trial was independently assessed by two reviewers (GLN and HCM), with disagreements resolved by a third author (BMD), using the Cochrane Risk of Bias 2 (RoB 2) tool, which comprises five domains: (1) the randomization process, (2) deviations from intended interventions, (3) missing outcome data, (4) measurement of the outcome, and (5) selection of the reported result [34].

### Certainty of the evidence

The certainty of the evidence for the main outcomes was assessed using the Grading of Recommendations Assessment, Development and Evaluation (GRADE) approach and rated as high, moderate, low, or very low [35]. For the primary outcomes, imprecision was judged against the prespecified MIDs.

### Use of AI

A large language model (Claude Opus 5.5, Anthropic) was used for linguistic revision and stylistic refinement to improve the clarity, readability, and overall quality of the manuscript in accordance with academic scientific writing standards. It was not used to generate or manipulate research data, perform statistical analyses, or draw scientific conclusions. All content was reviewed, edited, and validated by the authors, who take full responsibility for the manuscript.

## Results

### Study selection and characteristics

The systematic search identified 210 records. After removal of duplicates and application of the eligibility criteria, 23 reports were assessed in full text (Fig. 1). Three RCTs were ultimately included in the systematic review and meta-analysis [20–22]. The main reasons for exclusion were a study population without T2DM and the absence of a CagriSema 1.0/1.0 mg arm.

**Fig. 1.**
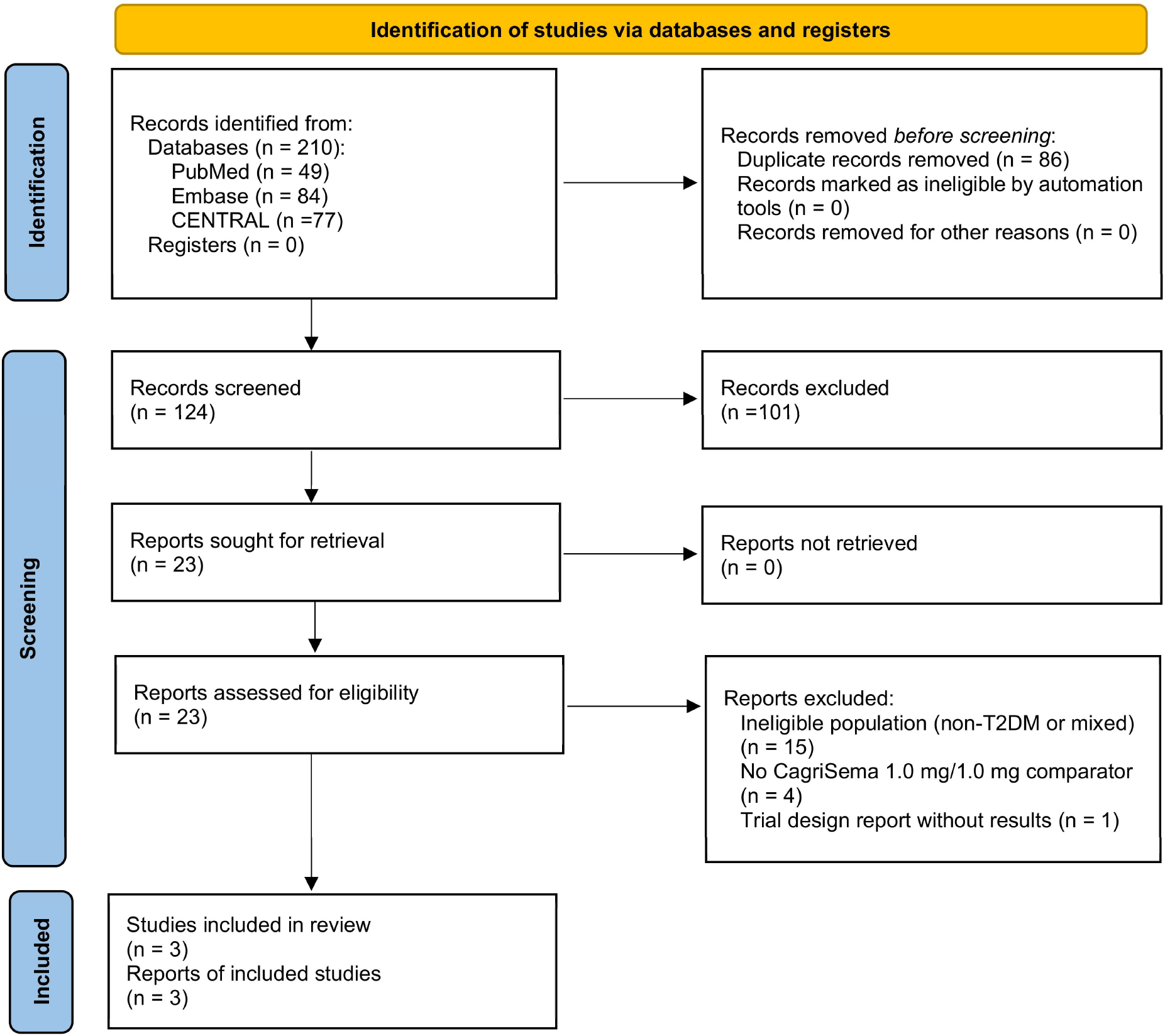
PRISMA 2020 flow diagram of study identification, screening, and inclusion. CENTRAL, Cochrane Central Register of Controlled Trials; PRISMA, Preferred Reporting Items for Systematic Reviews and Meta-Analyses; T2DM, type 2 diabetes mellitus

The three included trials randomized a total of 1,506 participants: 755 to CagriSema 2.4/2.4 mg and 751 to CagriSema 1.0/1.0 mg. Treatment duration was 40 or 68 weeks, and mean baseline HbA1c ranged from 7.7% (61 mmol/mol) to 8.8% (73 mmol/mol) across trials. Detailed study characteristics are presented in Table 1.

**Table 1.** Baseline characteristics of included studies.

| Study | Study design | Population | Study group | N | Age, years | Female, n (%) | HbA1c, % / mmol/mol | Body weight, kg | BMI, kg/m² | Waist circumference, cm | Duration of diabetes, years |
|---|---|---|---|---|---|---|---|---|---|---|---|
| Aroda, 2026 - REIMAGINE 1 | Multi-center RCT (40 wk), phase 3a | T2DM on diet and exercise; BMI ≥23 kg/m² | CagriSema 2.4/2.4 mg | 62 | 53.5 (12.6) | 32 (52) | 7.7 (0.7) / 60.7 (7.1) | 99.3 (23.5) | 35.3 (7.8) | 114.5 (16.8) | 2.7 (2.9) |
|  |  |  | CagriSema 1.0/1.0 mg | 63 | 53.3 (12.3) | 26 (41) | 7.9 (0.7) / 62.5 (7.4) | 100.9 (24.5) | 34.9 (7.5) | 115.1 (17.8) | 3.2 (4.2) |
| Buse, 2026 - REIMAGINE 2 | Multi-center RCT (68 wk), phase 3a | T2DM on metformin ± SGLT2i; BMI ≥25 kg/m² | CagriSema 2.4/2.4 mg | 603 | 57.2 (10.7) | 266 (44) | 8.2 (0.9) / 65.7 (10.2) | 99.2 (21.2) | 34.7 (6.5) | 114.1 (15.4) | 8.6 (6.5) |
|  |  |  | CagriSema 1.0/1.0 mg | 595 | 56.4 (10.6) | 250 (42) | 8.2 (1.0) / 66.4 (10.8) | 102.1 (23.9) | 35.4 (7.3) | 116.0 (17.0) | 8.4 (6.0) |
| Rosenstock, 2026 - REIMAGINE 3 | Multi-center RCT (40 wk), phase 3a | T2DM on basal insulin ± metformin; BMI ≥25 kg/m² | CagriSema 2.4/2.4 mg | 90 | 59.2 (9.3) | 38 (42) | 8.8 (0.9) / 72.7 (10.1) | 88.9 (18.5) | 32.0 (6.0) | 106.8 (16.0) | 15.1 (7.9) |
|  |  |  | CagriSema 1.0/1.0 mg | 93 | 58.8 (10.8) | 41 (44) | 8.8 (1.0) / 72.4 (10.5) | 86.7 (15.8) | 31.4 (5.5) | 104.9 (13.5) | 14.6 (7.4) |

**Note:** Data are presented as mean (SD) unless otherwise specified. CagriSema 2.4/2.4 mg consists of 2.4 mg cagrilintide combined with 2.4 mg semaglutide, and CagriSema 1.0/1.0 mg consists of 1.0 mg cagrilintide combined with 1.0 mg semaglutide. BMI, body mass index; HbA1c, glycated hemoglobin; RCT, randomized controlled trial; SD, standard deviation; SGLT2i, sodium–glucose cotransporter-2 inhibitor; T2DM, type 2 diabetes mellitus; wk, weeks; ±, with or without.

### Body weight

CagriSema 2.4/2.4 mg was associated with a significantly greater percentage reduction in body weight than CagriSema 1.0/1.0 mg (MD −2.07 pp; 95% CI −2.91 to −1.24; p < 0.001; I² = 0%; Fig. 2). As the entire 95% CI lay below the prespecified MID of 3 pp, the 1.0/1.0 mg dose was considered not meaningfully inferior for this outcome. The result was consistent in the sensitivity analysis using the treatment-policy estimand (MD −1.83 pp; 95% CI −2.66 to −0.99; Fig. S1).

**Fig. 2.**
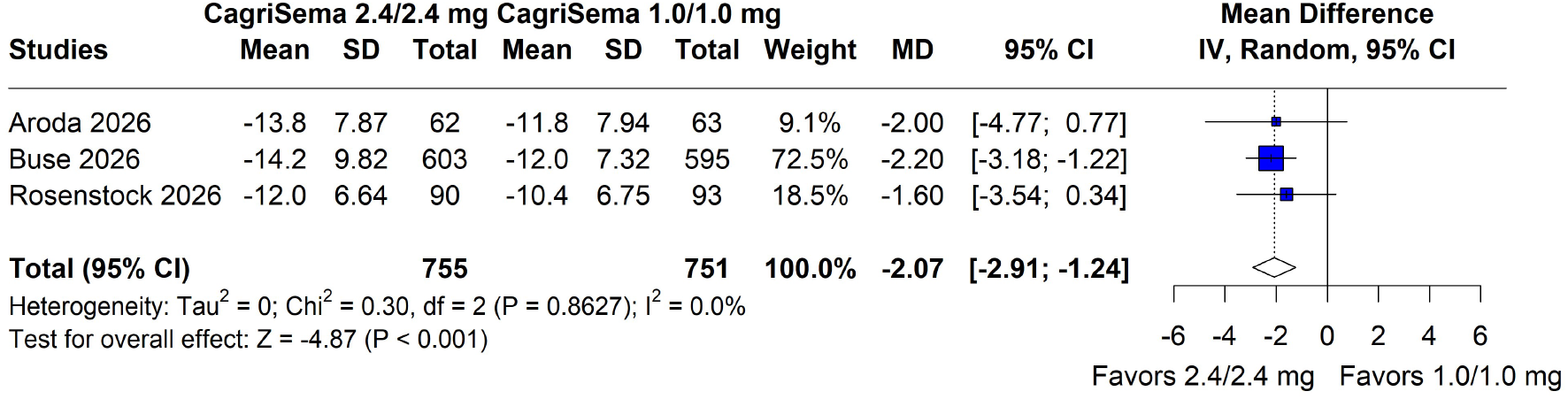
Percentage change in body weight: CagriSema 2.4/2.4 mg was associated with a significantly greater reduction than CagriSema 1.0/1.0 mg. Squares represent study-specific mean differences (size proportional to study weight), horizontal lines their 95% CIs, and the diamond the pooled random-effects estimate (MD −2.07 percentage points; 95% CI −2.91 to −1.24). CI, confidence interval; IV, inverse variance; MD, mean difference; SD, standard deviation

The 2.4/2.4 mg dose was also associated with higher proportions of participants achieving body-weight reductions of ≥10% (64.9% vs. 55.1%; RR 1.18; 95% CI 1.08 to 1.28; p < 0.001; I² = 0%; Fig. 3) and ≥15% (41.9% vs. 30.9%; RR 1.35; 95% CI 1.18 to 1.55; p < 0.001; I² = 0%; Fig. 4). There were no significant differences for reductions of ≥5% or ≥20% (Figs. S2 and S3).

**Fig. 3.**
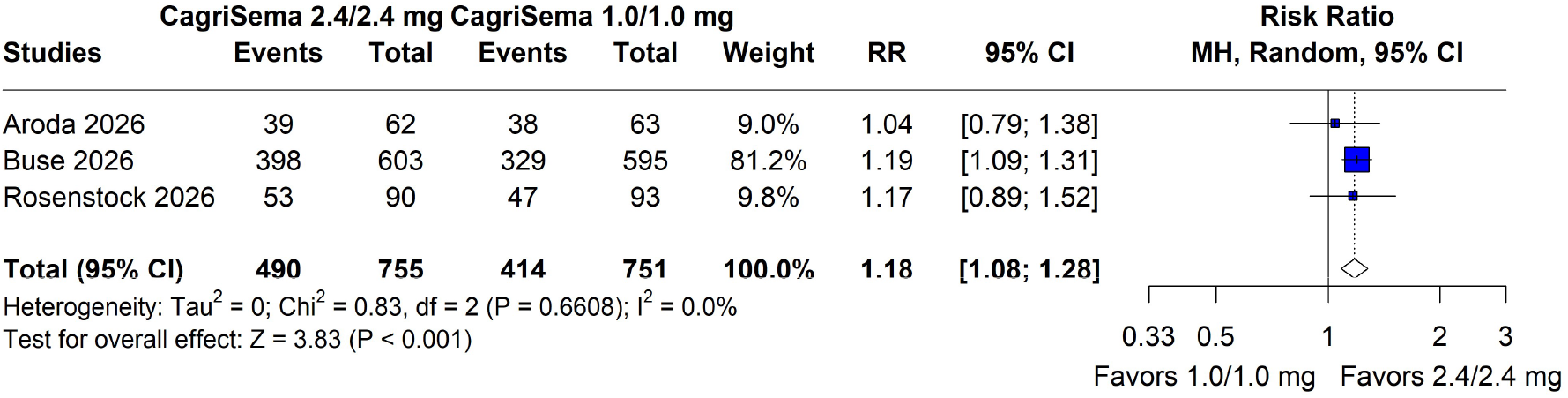
Proportion of participants achieving ≥10% body-weight reduction: CagriSema 2.4/2.4 mg was associated with a significantly higher proportion than CagriSema 1.0/1.0 mg. Squares represent study-specific risk ratios (size proportional to study weight), horizontal lines their 95% CIs, and the diamond the pooled random-effects estimate (RR 1.18; 95% CI 1.08 to 1.28). CI, confidence interval; MH, Mantel–Haenszel; RR, risk ratio

**Fig. 4.**
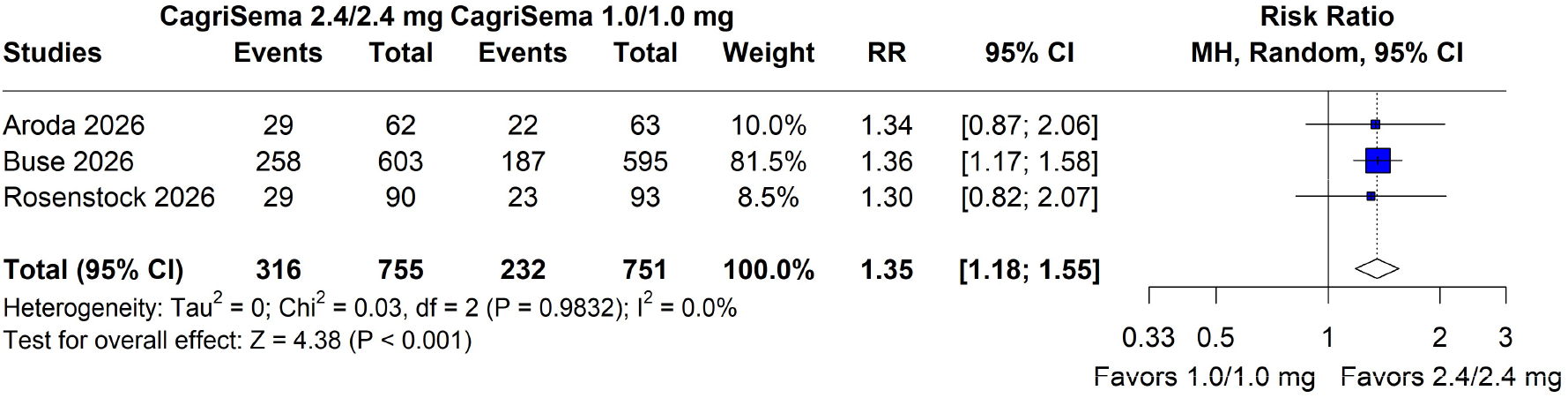
Proportion of participants achieving ≥15% body-weight reduction: CagriSema 2.4/2.4 mg was associated with a significantly higher proportion than CagriSema 1.0/1.0 mg. Squares represent study-specific risk ratios (size proportional to study weight), horizontal lines their 95% CIs, and the diamond the pooled random-effects estimate (RR 1.35; 95% CI 1.18 to 1.55). CI, confidence interval; MH, Mantel–Haenszel; RR, risk ratio

### Glycemic outcomes

In the primary analysis, the difference in HbA1c between doses was not statistically significant (MD −0.16 pp [−1.7 mmol/mol]; 95% CI −0.43 pp [−4.7 mmol/mol] to 0.10 [1.1 mmol/mol]; p = 0.118; I² = 35.8%; Fig. 5). However, as all three trials favored the 2.4/2.4 mg dose, a setting in which the Wald-type interval might be preferable, this method yielded a statistically significant difference (95% CI −0.29 [−3.2 mmol/mol] to −0.03 [−0.3 mmol/mol]; p = 0.016). Relative to the prespecified MID of 0.3 pp (3.3 mmol/mol), the result was inconclusive in the primary analysis, as its 95% CI included differences larger than the MID, whereas the entire Wald-type interval lay below the MID. The difference was also not statistically significant in the treatment-policy estimand analysis (MD −0.18 pp [−2.0 mmol/mol]; 95% CI −0.63 [−6.9 mmol/mol] to 0.26 [2.8 mmol/mol]; Fig. S4).

**Fig. 5.**
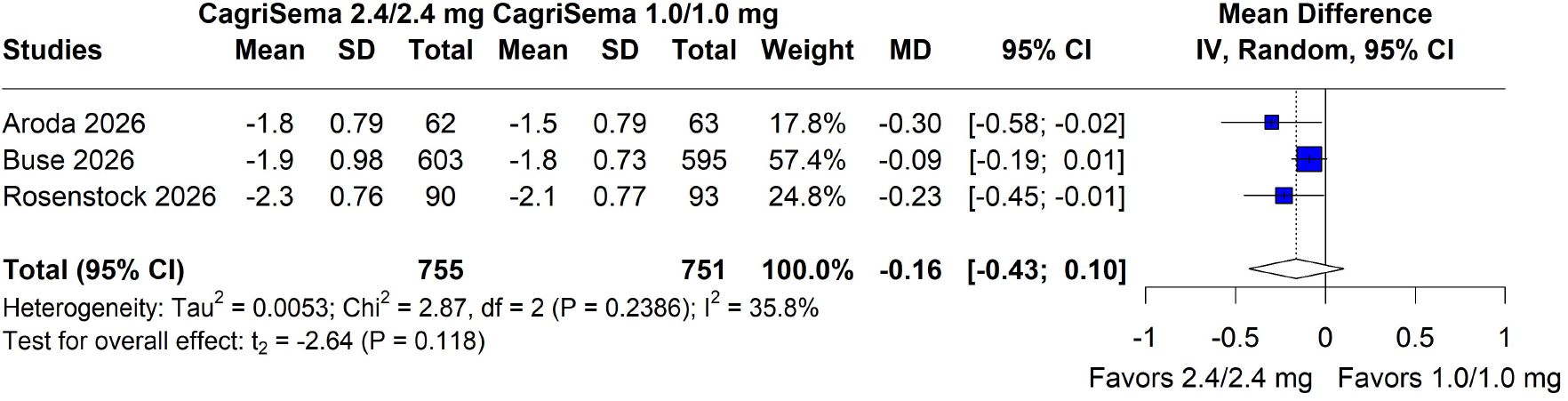
Change in HbA1c (percentage points): comparative analysis showed no statistically significant difference between CagriSema 2.4/2.4 mg and CagriSema 1.0/1.0 mg. Squares represent study-specific mean differences (size proportional to study weight), horizontal lines their 95% CIs, and the diamond the pooled random-effects estimate with HKSJ-adjusted CI (MD −0.16 percentage points [−1.7 mmol/mol]; 95% CI −0.43 to 0.10 [−4.7 to 1.1 mmol/mol]). Sensitivity analysis with a Wald-type CI: −0.29 to −0.03 (−3.2 to −0.3 mmol/mol). CI, confidence interval; HbA1c, glycated hemoglobin; HKSJ, Hartung–Knapp–Sidik–Jonkman; IV, inverse variance; MD, mean difference; SD, standard deviation

There was no significant difference in fasting plasma glucose (MD −0.22 mmol/L; 95% CI −0.72 to 0.27; p = 0.191; I² = 27.9%; Fig. 6). A slightly higher proportion of participants achieved HbA1c ≤6.5% (≤48 mmol/mol) with the 2.4/2.4 mg dose (RR 1.07; 95% CI 1.00 to 1.14; Fig. S5), with no significant differences for HbA1c <7.0% (<53 mmol/mol) or the use of glycemic rescue medication (Figs. S6 and S7).

**Fig. 6.**
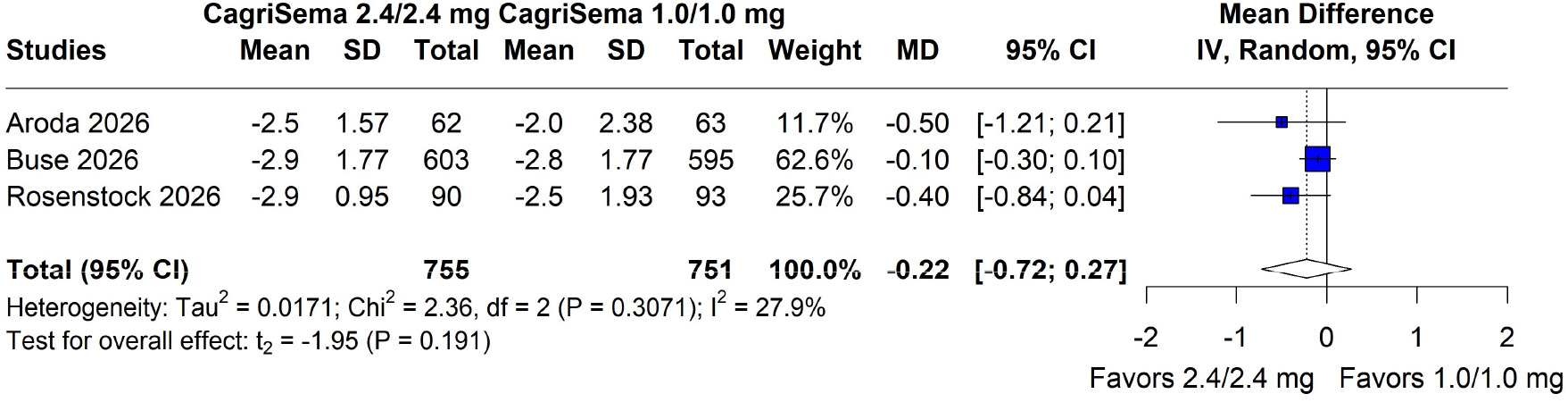
Change in fasting plasma glucose (mmol/L): comparative analysis showed no statistically significant difference between doses. Squares represent study-specific mean differences (size proportional to study weight), horizontal lines their 95% CIs, and the diamond the pooled random-effects estimate with HKSJ-adjusted CI (MD −0.22 mmol/L; 95% CI −0.72 to 0.27). Sensitivity analysis with a Wald-type CI: −0.48 to 0.03. CI, confidence interval; HKSJ, Hartung–Knapp–Sidik–Jonkman; IV, inverse variance; MD, mean difference; SD, standard deviation

### Waist circumference and blood pressure

CagriSema 2.4/2.4 mg was associated with a significantly greater reduction in waist circumference (MD −2.35 cm; 95% CI −4.29 to −0.41; p = 0.035; I² = 6.0%; Fig. 7). There was no significant difference in systolic blood pressure (MD −0.53 mmHg; 95% CI −1.77 to 0.72; p = 0.407; I² = 0%; Fig. 8) or diastolic blood pressure (Fig. S8).

**Fig. 7.**
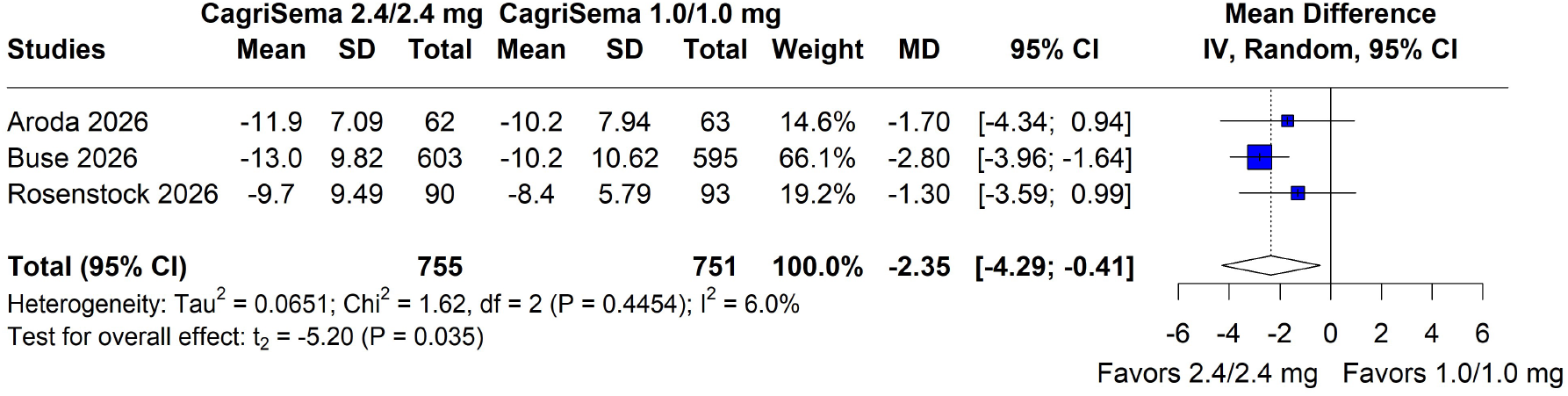
Change in waist circumference (cm): CagriSema 2.4/2.4 mg was associated with a significantly greater reduction than CagriSema 1.0/1.0 mg. Squares represent study-specific mean differences (size proportional to study weight), horizontal lines their 95% CIs, and the diamond the pooled random-effects estimate with HKSJ-adjusted CI (MD −2.35 cm; 95% CI −4.29 to −0.41). Sensitivity analysis with a Wald-type CI: −3.38 to −1.32. CI, confidence interval; HKSJ, Hartung–Knapp–Sidik–Jonkman; IV, inverse variance; MD, mean difference; SD, standard deviation

**Fig. 8.**
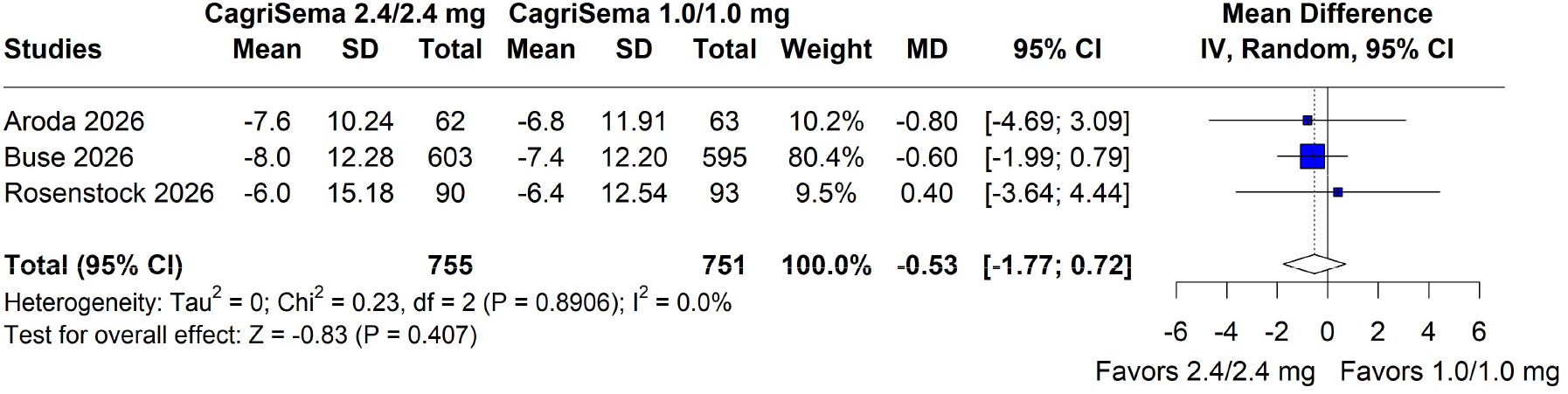
Change in systolic blood pressure (mmHg): comparative analysis showed no statistically significant difference between doses. Squares represent study-specific mean differences (size proportional to study weight), horizontal lines their 95% CIs, and the diamond the pooled random-effects estimate (MD −0.53 mmHg; 95% CI −1.77 to 0.72). CI, confidence interval; IV, inverse variance; MD, mean difference; SD, standard deviation

### Lipid profile and high-sensitivity C-reactive protein

There were no significant differences between doses in total cholesterol (RoM 0.99; 95% CI 0.93 to 1.06; p = 0.697; I² = 44.2%; Fig. 9) or LDL-C (RoM 1.00; 95% CI 0.90 to 1.11; p = 0.961; I² = 44.9%; Fig. 10), nor in non-HDL cholesterol or free fatty acids (Figs. S9 and S10). The 2.4/2.4 mg dose was associated with a slightly greater increase in high-density lipoprotein cholesterol (HDL-C; RoM 1.02; 95% CI 1.00 to 1.04; Fig. S11) and slightly greater reductions in triglycerides (RoM 0.96; 95% CI 0.92 to 0.99; Fig. S12) and very-low-density lipoprotein cholesterol (VLDL-C; RoM 0.96; 95% CI 0.92 to 0.99; Fig. S13). High-sensitivity C-reactive protein did not differ between doses (Fig. S14).

**Fig. 9.**
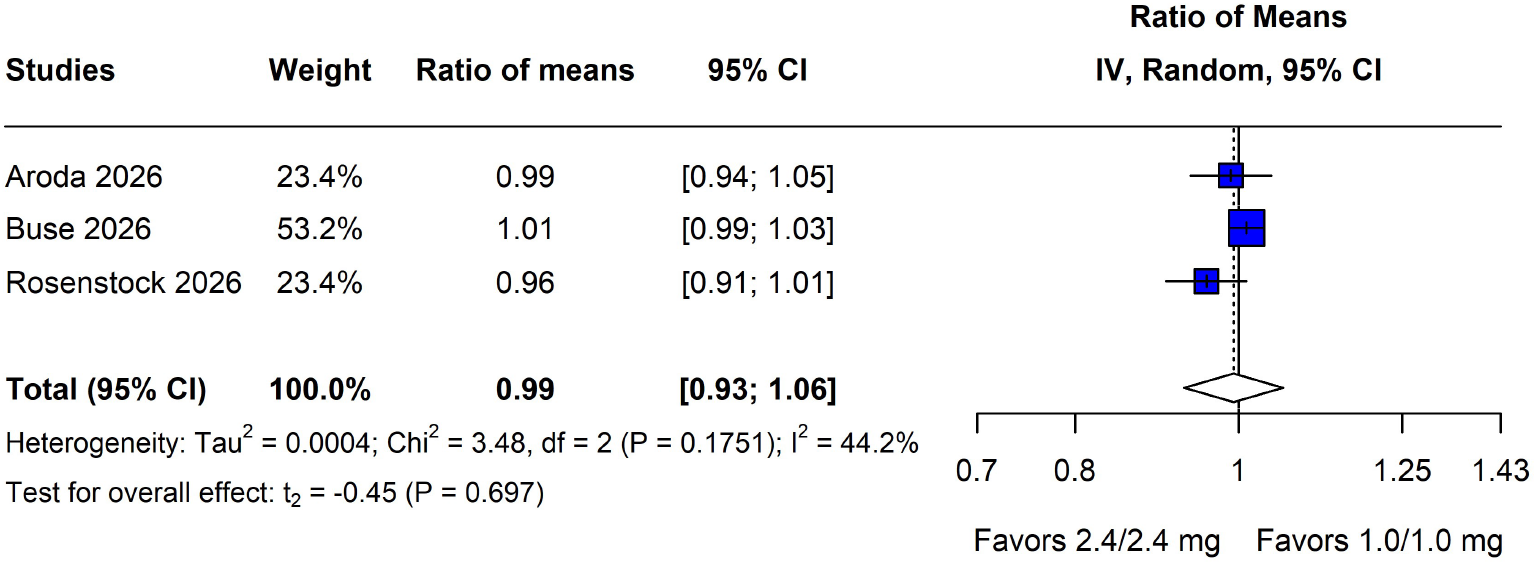
Total cholesterol (ratio to baseline): comparative analysis showed no statistically significant difference between doses. Squares represent study-specific ratios of geometric means (size proportional to study weight), horizontal lines their 95% CIs, and the diamond the pooled random-effects estimate with HKSJ-adjusted CI (RoM 0.99; 95% CI 0.93 to 1.06). Sensitivity analysis with a Wald-type CI: 0.96 to 1.03. CI, confidence interval; HKSJ, Hartung–Knapp–Sidik–Jonkman; IV, inverse variance; RoM, ratio of geometric means

**Fig. 10.**
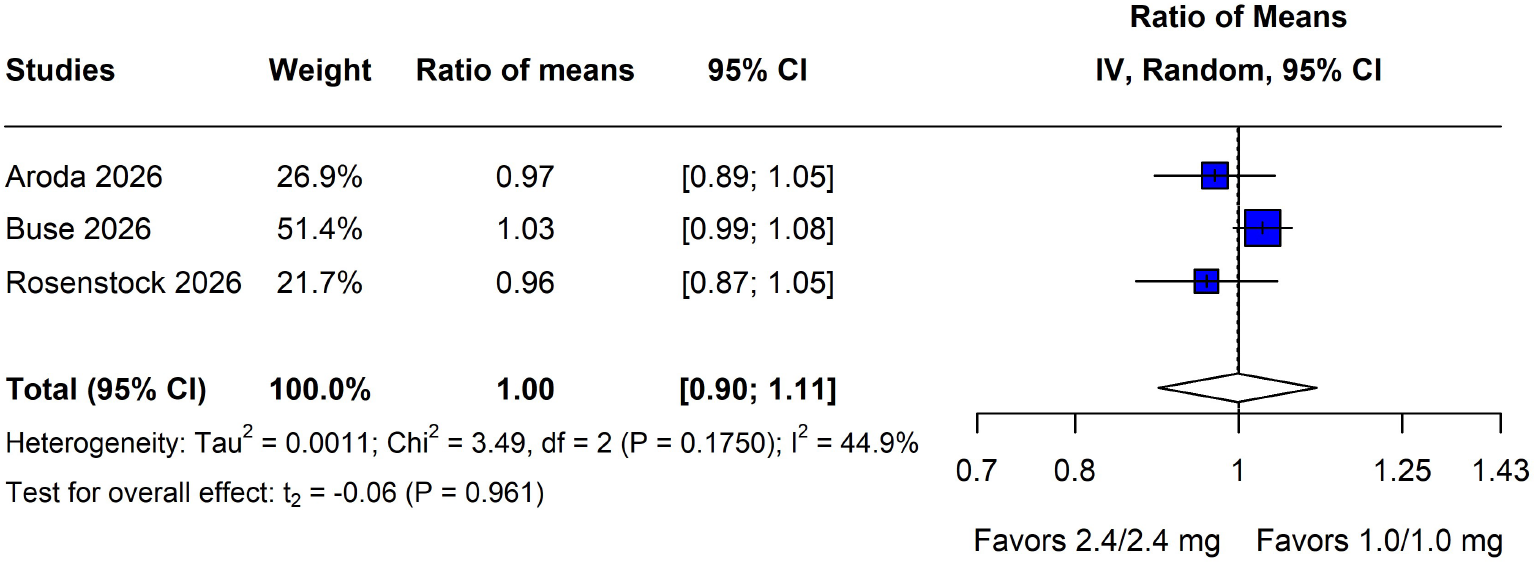
LDL cholesterol (ratio to baseline): comparative analysis showed no statistically significant difference between doses. Squares represent study-specific ratios of geometric means (size proportional to study weight), horizontal lines their 95% CIs, and the diamond the pooled random-effects estimate with HKSJ-adjusted CI (RoM 1.00; 95% CI 0.90 to 1.11). Sensitivity analysis with a Wald-type CI: 0.95 to 1.05. CI, confidence interval; HKSJ, Hartung–Knapp–Sidik–Jonkman; IV, inverse variance; LDL, low-density lipoprotein; RoM, ratio of geometric means

### Quality of life and treatment satisfaction

There were no significant differences between doses in the 36-Item Short Form Health Survey version 2 (SF-36v2) physical or mental component summary scores or in the Diabetes Treatment Satisfaction Questionnaire (DTSQ) score (Figs. S15–S17).

### Safety and tolerability

The incidence of any adverse event was slightly higher with CagriSema 2.4/2.4 mg (85.4% vs. 79.7%; RR 1.07; 95% CI 1.02 to 1.12; p = 0.004; I² = 0%; Fig. 11), as was that of gastrointestinal disorders (64.8% vs. 53.2%; RR 1.22; 95% CI 1.12 to 1.32; p < 0.001; I² = 0%; Fig. 12). There were no significant differences in serious adverse events (9.7% vs. 9.9%; RR 0.98; 95% CI 0.72 to 1.33; p = 0.890; I² = 0%; Fig. 13) or adverse events leading to permanent treatment discontinuation (7.8% vs. 7.3%; RR 0.98; 95% CI 0.37 to 2.61; p = 0.930; I² = 25.7%; Fig. 14), including those of gastrointestinal nature (Fig. S18).

**Fig. 11.**
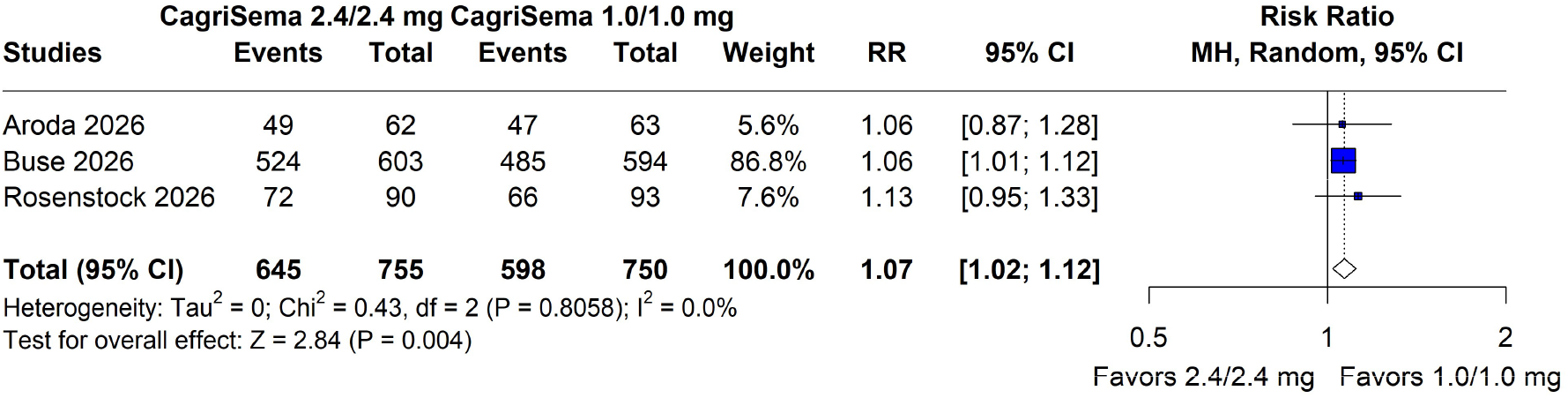
Any adverse event: CagriSema 2.4/2.4 mg was associated with a significantly higher incidence than CagriSema 1.0/1.0 mg. Squares represent study-specific risk ratios (size proportional to study weight), horizontal lines their 95% CIs, and the diamond the pooled random-effects estimate (RR 1.07; 95% CI 1.02 to 1.12). CI, confidence interval; MH, Mantel–Haenszel; RR, risk ratio

**Fig. 12.**
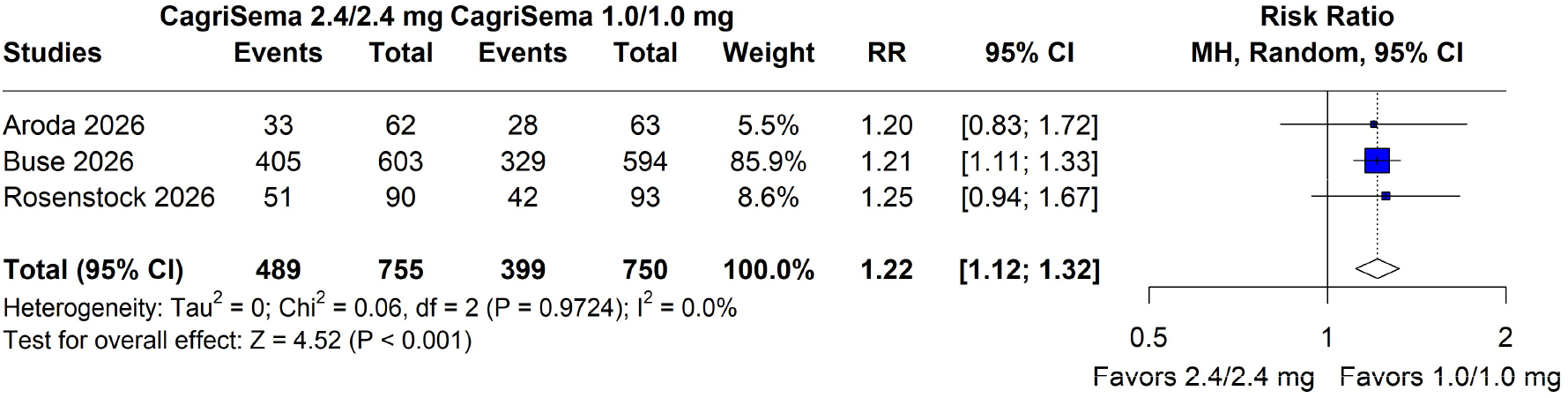
Gastrointestinal disorders: CagriSema 2.4/2.4 mg was associated with a significantly higher incidence than CagriSema 1.0/1.0 mg. Squares represent study-specific risk ratios (size proportional to study weight), horizontal lines their 95% CIs, and the diamond the pooled random-effects estimate (RR 1.22; 95% CI 1.12 to 1.32). CI, confidence interval; MH, Mantel–Haenszel; RR, risk ratio

**Fig. 13.**
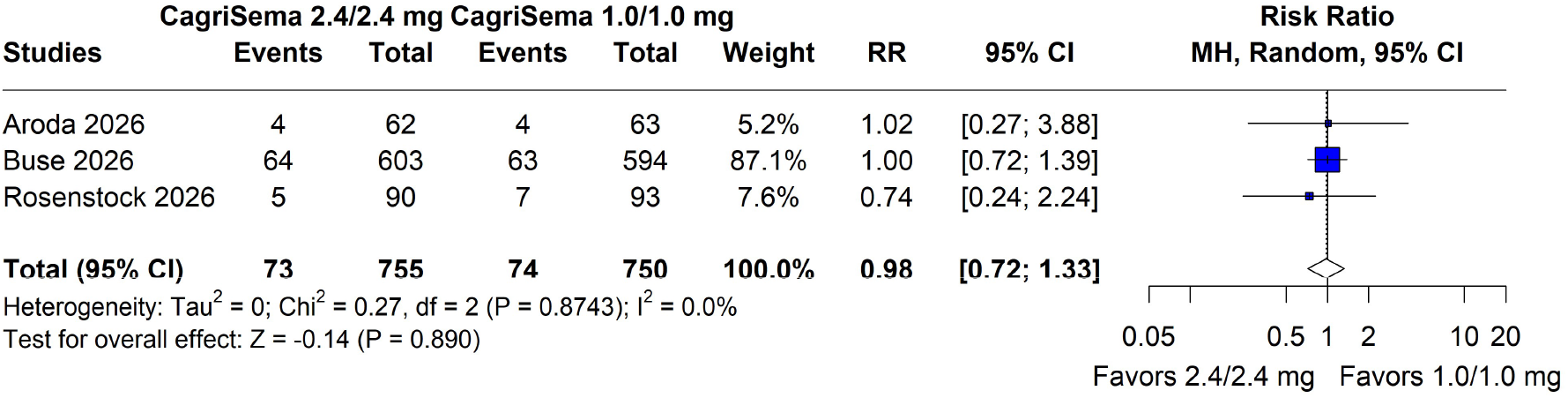
Serious adverse events: comparative analysis showed no statistically significant difference between doses. Squares represent study-specific risk ratios (size proportional to study weight), horizontal lines their 95% CIs, and the diamond the pooled random-effects estimate (RR 0.98; 95% CI 0.72 to 1.33). CI, confidence interval; MH, Mantel–Haenszel; RR, risk ratio

**Fig. 14.**
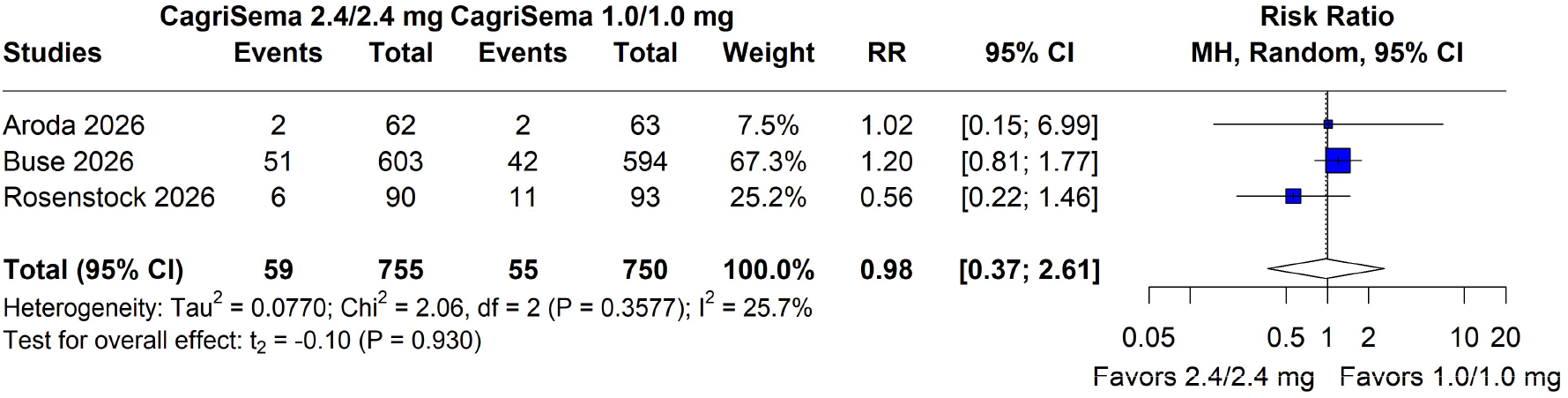
Adverse events leading to permanent treatment discontinuation: comparative analysis showed no statistically significant difference between doses. Squares represent study-specific risk ratios (size proportional to study weight), horizontal lines their 95% CIs, and the diamond the pooled random-effects estimate with HKSJ-adjusted CI (RR 0.98; 95% CI 0.37 to 2.61). Sensitivity analysis with a Wald-type CI: 0.56 to 1.69. CI, confidence interval; HKSJ, Hartung–Knapp–Sidik–Jonkman; MH, Mantel–Haenszel; RR, risk ratio Among individual adverse events, the 2.4/2.4 mg dose was associated with a higher incidence of nausea (RR 1.29; 95% CI 1.11 to 1.50; Fig. S19), vomiting (RR 1.34; 95% CI 1.04 to 1.74; Fig. S20), decreased appetite (RR 1.47; 95% CI 1.14 to 1.90; Fig. S21), and fatigue (RR 1.58; 95% CI 1.11 to 2.24; Fig. S22). There were no significant differences in diarrhea, constipation, injection-site reactions, level 1, 2, or 3 hypoglycemia, retinal disorders, gallbladder-related disorders, neoplasms, malignant neoplasms, hypersensitivity reactions, or deaths (Figs. S23–S34).

Fewer participants reached the assigned target maintenance dose with the 2.4/2.4 mg dose, although the difference was not statistically significant in the primary analysis (RR 0.84; 95% CI 0.64 to 1.09; Fig. S35); however, with all three trials pointing in the same direction, the Wald-type method yielded a statistically significant difference (95% CI 0.74 to 0.95). Premature treatment discontinuation for any reason did not differ between doses (Fig. S36).

### Risk-of-bias assessment

Using the RoB 2 tool, all three included studies were judged to be at overall low risk of bias across all five domains assessed (Supplementary File 2, Table S2).

### Certainty of the evidence

According to the GRADE assessment, the certainty of evidence was high for percentage change in body weight, waist circumference, proportion of participants achieving ≥10% and ≥15% body-weight reduction, any adverse event and any gastrointestinal event. Certainty was moderate for the remaining main outcomes, primarily reflecting imprecision. Full GRADE judgments are presented in Supplementary File 2 (Table S3).

## Discussion

To our knowledge, this is the first systematic review and meta-analysis to compare CagriSema 2.4/2.4 mg with CagriSema 1.0/1.0 mg in adults with T2DM, including three RCTs and 1,506 participants. The main findings were as follows: (1) the 2.4/2.4 mg dose was associated with a statistically significant additional reduction in body weight of 2.07 pp, although the entire 95% CI of this difference lay below the prespecified MID of 3 pp; (2) there was no statistically significant difference in HbA1c in the primary analysis; (3) differences in the remaining cardiometabolic parameters were absent or of small magnitude; and (4) the 2.4/2.4 mg dose was associated with a higher incidence of any adverse event and of gastrointestinal disorders, without an increase in serious adverse events or in adverse events leading to treatment discontinuation.

Our findings complement previous meta-analyses showing the benefit of CagriSema over semaglutide or cagrilintide monotherapy and placebo [10, 23, 28]. In adults with overweight or obesity, with or without T2DM, Shao et al. reported an additional reduction in body weight of 6.6 pp with CagriSema 2.4/2.4 mg compared with semaglutide 2.4 mg [10]. In a recent network meta-analysis of 262 trials, CagriSema was among the most effective agents, with an estimated weight reduction of 14.8% versus lifestyle modification alone at one year, second only to tirzepatide (14.9%) [36]. However, previous syntheses either focused on the 2.4/2.4 mg dose or compared each dose separately with other comparators [23]. By pooling the within-trial randomized comparison between doses, the present review shows that the 1.0/1.0 mg dose also produced substantial metabolic effects, with 55% of participants achieving a body-weight reduction of at least 10%.

The additional weight reduction of 2.07 pp with the 2.4/2.4 mg dose was smaller than the prespecified MID of 3 pp. This MID was estimated based on STEP 2, in which semaglutide 2.4 mg reduced body weight by 3.1 pp more than semaglutide 1.0 mg in adults with T2DM (trial product estimand: −10.6% vs. −7.6%) [29]. Because escalation of CagriSema increases the doses of both components, a similar or larger increment was considered plausible, and no established non-inferiority margin exists for weight-loss outcomes [37]. One possible explanation for the modest additional weight reduction could be attributed to the substantial weight loss attained with the 1.0/1.0 mg dose (10.4% to 12.0% across trials), which may indicate a flattening of the dose–response curve. However, this cross-trial comparison should be interpreted cautiously given differences in populations and treatment duration, and the fact that the entire CI lay below the MID does not constitute formal evidence of non-inferiority.

For HbA1c, the difference between doses was also small but was not statistically significant in the primary analysis, and its CI did not exclude a clinically relevant difference in favor of the 2.4/2.4 mg dose, rendering the findings inconclusive regarding the MID. This is consistent with semaglutide dose-escalation trials, in which higher doses produced additional weight loss but only modest additional HbA1c reductions [16, 29]. Both doses produced large HbA1c reductions (1.5 to 2.3 pp [16.4 to 25.1 mmol/mol] across trials), and a slightly higher proportion of participants achieved HbA1c ≤6.5% (≤48 mmol/mol) with the higher dose. Together, these findings suggest that escalation to the 2.4/2.4 mg dose adds more to weight loss than to glycemic control.

The 2.4/2.4 mg dose was associated with a higher incidence of gastrointestinal disorders (64.8% vs. 53.2%), particularly nausea and vomiting, without differences in serious adverse events or treatment discontinuation. Dose-dependent gastrointestinal effects have also been described for other incretin-based therapies, such as tirzepatide [38]. In all three included trials, dose escalation could be delayed or the dose reduced in the case of intolerable adverse events, and participants could remain on the maximum tolerated dose [20–22]. Although flexible dosing regimens may have supported treatment adherence, only 73.6% of those assigned to 2.4/2.4 mg reached the target dose at any time, compared with 92.1% in the 1.0/1.0 mg group. The REIMAGINE 2 investigators suggested that gastrointestinal intolerance, the substantial weight loss already achieved, and reluctance among participants or investigators to escalate may have contributed [22]. Remaining on lower doses may support individualized treatment, but it also attenuates the contrast in efficacy between the assigned regimens.

The 2026 American Diabetes Association and the European Association for the Study of Diabetes consensus report places individualized weight management alongside glycemic control as a core goal in the management of T2DM [39]. At the time of writing, CagriSema has not been approved by the FDA; an application for chronic weight management at the 2.4/2.4 mg dose is under review [40], and no dose has been established for the treatment of T2DM. In this context, the additional weight loss with the 2.4/2.4 mg dose should be weighed against the absence of a clear additional HbA1c benefit, the higher incidence of gastrointestinal events, and the lower proportion of participants reaching the target dose. The 1.0/1.0 mg dose may therefore be a reasonable option when glycemic control is the main goal or gastrointestinal tolerability is a concern, whereas the 2.4/2.4 mg dose may be preferred when additional weight loss is a priority.

This study has several limitations. First, only three trials were available, one of which contributed approximately 80% of the participants and most of the statistical weight in the pooled analyses. Second, the small number of studies precluded leave-one-out sensitivity analyses, meta-regression, and formal assessment of publication bias. Third, no trial reported a direct comparison between the two doses, and study-level contrasts were therefore constructed from the adjusted arm-level estimates, assuming independence between them; although this approximation adequately reproduced the published SEs of other contrasts within the same trials, it may not exactly recover the precision of the between-dose comparison. Finally, the trials differed in background therapy, stage of T2DM, and treatment duration; statistical heterogeneity was nevertheless absent or low for most outcomes, and inconsistency was considered in the GRADE assessment.

## Conclusion

In adults with T2DM, CagriSema 2.4/2.4 mg produced greater weight loss than 1.0/1.0 mg, but the difference was smaller than the prespecified MID; there was no clear additional benefit on HbA1c, and gastrointestinal adverse events were more frequent. These findings support individualized dose selection, with the 1.0/1.0 mg dose as a reasonable option when glycemic control, rather than additional weight loss, is the main treatment goal.

## Statements and Declarations

### Ethics approval

Not applicable. This systematic review and meta-analysis used aggregate data from published studies, each of which reported approval by the relevant ethics committees.

### Consent to participate

Not applicable.

### Consent to publish

Not applicable.

### Data availability

All data analyzed in this study were extracted from published studies cited in the article; the extraction datasets and analytic code are available from the corresponding author upon reasonable request.

### Author contributions

B.M.D.: conceptualization (lead), methodology (lead), investigation (lead), data curation (lead), formal analysis (lead), visualization (lead), project administration (lead), writing – original draft (lead). L.E.C.: formal analysis (supporting), writing – original draft (supporting). G.M.P.: formal analysis (supporting), writing – original draft (supporting). H.C.M.: formal analysis (supporting). G.L.N.: investigation (supporting), data curation (supporting), formal analysis (supporting). L.O.M.: data curation (supporting), visualization (supporting). G.G.S.: investigation (supporting), data curation (supporting). F.G.: conceptualization (supporting), supervision (lead), writing - review and editing (lead). All authors critically revised the manuscript for important intellectual content, approved the final version, and agreed to be accountable for all aspects of the work.

## Acknowledgments

None.

## Supporting Information

**Supplementary File 1.** PRISMA 2020 checklist.

**Supplementary File 2.** Supplementary methods, search strategies, risk-of-bias assessment, GRADE evidence profile, and supplementary forest plots

