## Supplementary File 2 for "CagriSema 2.4/2.4 mg versus CagriSema 1.0/1.0 mg in adults with type 2 diabetes: a dose-comparison systematic review and meta-analysis of randomized controlled trials"

**Supplementary Methods S1**

Data sources, estimands, and analysis populations

All three included trials reported efficacy outcomes under two estimands. Data from the efficacy (trial product) estimand, which estimates the treatment effect had all participants remained on the randomized treatment without rescue medication, were used for all efficacy analyses, as prespecified in the protocol and in accordance with previous meta-analyses on this topic [10 - reference numbers refer to the reference list of the main article]. Data from the treatment-policy estimand, which uses all available data regardless of treatment discontinuation or rescue medication, were used only in the sensitivity analyses of the primary outcomes.

For efficacy outcomes, the number of randomized participants per arm was used as the sample size (62 vs. 63, 603 vs. 595, and 90 vs. 93 participants in the 2.4/2.4 mg and 1.0/1.0 mg arms, respectively), because the published model-based estimates refer to the full randomized population, with missing data handled by the trials' statistical models. For safety outcomes, the safety analysis set reported by each trial (randomized participants who received at least one dose of trial product) was used; it coincided with the randomized population in all arms except the CagriSema 1.0/1.0 mg arm of REIMAGINE 2 (594 vs. 595 participants).

Continuous outcomes: contrasts from arm-level adjusted means

None of the trials reported a direct comparison between the two CagriSema doses. Each trial reported, for every arm, the estimated mean change from baseline with its SE, obtained from mixed models for repeated measures (MMRM) adjusted for the baseline value of the outcome and other covariates, together with estimated treatment differences of each CagriSema dose versus placebo or versus the corresponding semaglutide dose. The study-level contrast between doses was therefore obtained as:

$$MD = \text{mean}(2.4/2.4 \text{ mg}) - \text{mean}(1.0/1.0 \text{ mg}) \quad (1)$$

$$Var(MD) = SE(2.4)^2 + SE(1.0)^2 - 2 \cdot Cov(2.4, 1.0) \quad (2)$$

Because the covariance between the two adjusted means was not reported, it was assumed to be zero, so that $SE(MD) = \sqrt{[SE(2.4)^2 + SE(1.0)^2]}$. For simple sample means from independent randomized groups, this covariance is zero by definition [25]. For adjusted means estimated jointly by the same model, the covariance is not necessarily zero; however, with adequate randomization, similarly sized arms, and adjustment for the same covariates, the uncertainty shared by the two estimates is expected to be small, and the assumption was considered a reasonable approximation.

To allow pooling from arm-level summary data, each arm-level SE was converted to an effective SD:

$$SD = SE \times \sqrt{N} \quad (3)$$

N is the number of randomized participants in the arm [25]. Because the SE derives from an adjusted model rather than from raw sample means, this SD does not represent the observed dispersion of individual values; it was therefore not used to calculate standardized mean differences. In the inverse-variance calculation of the MD, the variance of each arm is recovered as $SD^2/N = SE^2$, so the effective SD reproduces the original arm-level SE and alters neither the point estimate nor its precision. The resulting SE(MD) is identical to that obtained from equation (2).

<u>Empirical check of the independence assumption</u>

The adequacy of the zero-covariance assumption was assessed using the trials' own data of the outcomes analyzed in the present review involving a CagriSema arm. For every contrast in which a trial reported both the arm-level SEs and the 95% CI of the corresponding estimated treatment difference (each CagriSema dose versus placebo in REIMAGINE 1 and 3, and versus the corresponding semaglutide or cagrilintide dose in REIMAGINE 2), two SEs were obtained: the SE reconstructed from the arm-level SEs under independence, SE(rec) = $\sqrt{[SE(arm A)^2 + SE(arm B)^2]}$, and the SE derived from the published CI, SE(pub) = (upper limit − lower limit)/3.92. The ratio SE(rec)/SE(pub) was then calculated for each contrast.

Across the 146 contrasts evaluated (136 versus placebo in REIMAGINE 1 and 3; 8 versus semaglutide and 2 versus cagrilintide in REIMAGINE 2), covering the outcomes and estimands for which both pieces of information were available, the median ratio was 1.00, and 92% of the ratios lay within ±10% of unity. The remaining discrepancies were small and compatible with the rounding of the published arm-level SEs, which were frequently reported to one or two decimal places. These findings indicate that, in these trials, ignoring the covariance between arm-level adjusted means had a negligible impact and that the independence assumption adequately reproduced the precision of the published treatment contrasts.

Arms without a reported SE

For several outcomes in REIMAGINE 2 (fasting plasma glucose, diastolic blood pressure, lipid parameters, high-sensitivity C-reactive protein, SF-36v2 component scores, DTSQ score, and waist circumference in the 1.0/1.0 mg arm), as well as for its treatment-policy estimand data, only the arm-level mean was reported, together with the 95% CI of the estimated treatment difference versus the corresponding semaglutide dose (605 participants randomized to semaglutide 2.4 mg and 609 to semaglutide 1.0 mg). In these cases, the SE of the difference was first obtained from the CI, and the arm-level SE of CagriSema was then derived assuming equal SDs in the CagriSema and semaglutide arms [25]:

SE(difference) = (upper limit − lower limit)/3.92 (4)

$SD(common) = SE(difference)/\sqrt{(1/N\_CagriSema + 1/N\_semaglutide)}$ (5)

$SE(CagriSema\ arm) = SD(common)/\sqrt{N\_CagriSema}$ (6)

Equation (5) relies on the same independence assumption described in “Continuous outcomes: contrasts from arm-level adjusted means”, applied in reverse. Because the published CI derives from an adjusted contrast rather than from a difference between raw means, this derivation is an algebraic approximation and does not recover the actual participant-level SDs. Its adequacy was checked in the six REIMAGINE 2 contrasts for which both arm-level SEs were published (CagriSema 2.4/2.4 mg versus semaglutide 2.4 mg and CagriSema 1.0/1.0 mg versus semaglutide 1.0 mg): in all six, the arm-level SE derived with equations (4)–(6) fell within the rounding range of the published arm-level SEs. The protocol had anticipated dividing the SE of the difference equally between the two arms; the formula above was used instead because it accounts for unequal arm sizes, and the two approaches yielded arm-level SEs differing by less than 0.5%.

Ratios to baseline

Lipid parameters and high-sensitivity C-reactive protein were reported as the geometric mean of the ratio of the end-of-treatment to the baseline value. Because the ratio is dimensionless, differences in reporting units across trials (mmol/L or mg/dL) did not affect the analysis. The analyses were performed on the natural log scale:

$$TE = \ln(\text{ratio } 2.4/2.4 \text{ mg}) - \ln(\text{ratio } 1.0/1.0 \text{ mg}) \quad (7)$$

$$SE(TE) = \sqrt{[\ln(SE\times\ 2.4)^2 + \ln(SE\times\ 1.0)^2]} \quad (8)$$

SE× denotes the multiplicative SE reported for each arm (REIMAGINE 1 and 3). In REIMAGINE 2, the multiplicative SE was either not reported or, for triglycerides, non-HDL cholesterol, and high-sensitivity C-reactive protein, was reported rounded to one decimal place as 1.0, which would imply a log-scale SE of zero and an infinite study weight. For this trial, the log-scale SE of each arm was therefore derived from the 95% CI of the estimated treatment ratio versus the corresponding semaglutide dose, applying equations (4)–(6) on the log scale; the reported geometric mean ratios themselves were retained. Pooled estimates were back-transformed and expressed as the ratio of geometric means (RoM); values <1 indicate a lower end-of-treatment-to-baseline ratio with CagriSema 2.4/2.4 mg.

Binary efficacy outcomes from model-estimated proportions

The proportions of participants achieving body-weight reductions of ≥5%, ≥10%, ≥15%, and ≥20%, and HbA1c targets of <7.0% and ≤6.5%, were reported as model-estimated proportions without event counts. The number of events in each arm was approximated as:

events = round(proportion/100 × N randomized) (9)

The number of randomized participants was used as the denominator because these proportions are estimated for the full randomized population, with outcomes imputed by the trials' models for participants without an end-of-treatment measurement.

**Supplementary Table S1.** Search strategy in each database.

| | |
|---|---|
| PubMed | (((cagrilintide OR “amylin analogue” OR “amylin analogues” OR “amylin analog” OR “amylin analogs” OR “amylin receptor agonist” OR AM833 OR NN9838 OR "cagrilintide" [Supplementary Concept]) AND (semaglutide OR Ozempic OR Wegovy OR Rybelsus OR NN9535 OR "Semaglutide"[Mesh])) OR (CagriSema OR NN9388)) AND (randomized controlled trial[pt] OR controlled clinical trial[pt] OR clinical trials as topic[mesh:noexp] OR trial[ti] OR random*[tiab] OR placebo*[tiab]) |
| Embase | (('cagrilintide' OR 'amylin analogue' OR 'amylin analogues' OR 'amylin analog' OR 'amylin analogs' OR 'amylin receptor agonist' OR 'am833' OR 'nn9838') AND ('semaglutide' OR 'ozempic' OR 'wegovy' OR 'rybelsus' OR 'nn9535' OR 'semaglutide'/exp) OR 'cagrisema' OR 'nn9388') AND ('controlled clinical trial'/exp OR random*:ti,ab OR placebo*:ti,ab OR trial:ti) AND [embase]/lim |
| Cochrane CENTRAL | (((cagrilintide OR “amylin analogue” OR “amylin analogues” OR “amylin analog” OR “amylin analogs” OR “amylin receptor agonist” OR AM833 OR NN9838) AND (semaglutide OR Ozempic OR Wegovy OR Rybelsus OR NN9535)) OR (CagriSema OR NN9388)) |

**Supplementary Table S2**. Risk of bias assessment across the included studies using the RoB 2.0 tool.

Risk of bias domains

| Study | D1 | D2 | D3 | D4 | D5 | Overall |
|---|---|---|---|---|---|---|
| Aroda 2026 | + | + | + | + | + | + |
| Buse 2026 | + | + | + | + | + | + |
| Rosenstock 2026 | + | + | + | + | + | + |

Domains:
D1: Bias arising from the randomization process.
D2: Bias due to deviations from intended intervention.
D3: Bias due to missing outcome data.
D4: Bias in measurement of the outcome.
D5: Bias in selection of the reported result.

Judgement
+ Low

**Supplementary Table S3.** GRADE assessment.

| Certainty assessment | | | | | | № of patients | | Effect | | Certainty | Importance |
|---|---|---|---|---|---|---|---|---|---|---|---|
| **№ of studies** | **Study design** | **Risk of bias** | **Inconsistency** | **Indirectness** | **Imprecision** | **CagriSema 2.4 mg / 2.4 mg** | **CagriSema 1.0 mg / 1.0 mg** | **Relative (95% CI)** | **Absolute (95% CI)** | | |
| **Percentage change in body weight** | | | | | | | | | | | |
| 3 | RCTs | not serious | not serious | not serious | not serious | 755 | 751 | - | MD **2.07 percentage points lower** (2.91 lower to 1.24 lower) | ⊕⊕⊕⊕ High | CRITICAL |
| **Change in HbA1c** | | | | | | | | | | | |
| 3 | RCTs | not serious | not serious | not serious | serious[a] | 755 | 751 | - | MD **0.16 percentage points lower** (0.43 lower to 0.10 higher) | ⊕⊕⊕◯ Moderate[a] | CRITICAL |
| **Fasting plasma glucose** | | | | | | | | | | | |
| 3 | RCTs | not serious | not serious | not serious | serious[b] | 755 | 751 | - | MD **0.22 mmol/L lower** (0.72 lower to 0.27 higher) | ⊕⊕⊕◯ Moderate[b] | IMPORTANT |
| **Waist circumference** | | | | | | | | | | | |
| 3 | RCTs | not serious | not serious | not serious | not serious | 755 | 751 | - | MD **2.35 cm lower** (4.29 lower to 0.41 lower) | ⊕⊕⊕⊕ High | IMPORTANT |
| **Systolic blood pressure** | | | | | | | | | | | |
| 3 | RCTs | not serious | not serious | not serious | serious[b] | 755 | 751 | - | MD **0.53 mmHg lower** (1.77 lower to 0.72 higher) | ⊕⊕⊕◯ Moderate[b] | IMPORTANT |
| **Total cholesterol (ratio to baseline)** | | | | | | | | | | | |
| 3 | RCTs | not serious | not serious | not serious | serious[b] | 755 | 751 | - | RoM **0.99** (0.93 to 1.06) | ⊕⊕⊕◯ Moderate[b] | IMPORTANT |
| **LDL cholesterol (ratio to baseline)** | | | | | | | | | | | |
| 3 | RCTs | not serious | not serious | not serious | serious[b] | 755 | 751 | - | RoM **1.00** (0.90 to 1.11) | ⊕⊕⊕◯ Moderate[b] | IMPORTANT |

| | | | | | | | | | | | |
|---|---|---|---|---|---|---|---|---|---|---|---|
| **Proportions of participants achieving ≥10% body-weight reduction** | | | | | | | | | | | |
| 3 | RCTs | not serious | not serious | not serious | not serious | 490/755 (64.9%) | 414/751 (55.1%) | **RR 1.18** (1.08 to 1.28) | **99 more per 1,000** (from 44 more to 154 more) | ⊕⊕⊕⊕ High | IMPORTANT |
| **Proportions of participants achieving ≥15% body-weight reduction** | | | | | | | | | | | |
| 3 | RCTs | not serious | not serious | not serious | not serious | 316/755 (41.9%) | 232/751 (30.9%) | **RR 1.35** (1.18 to 1.55) | **108 more per 1,000** (from 56 more to 170 more) | ⊕⊕⊕⊕ High | IMPORTANT |
| **Any adverse event** | | | | | | | | | | | |
| 3 | RCTs | not serious | not serious | not serious | not serious | 645/755 (85.4%) | 598/750 (79.7%) | **RR 1.07** (1.02 to 1.12) | **56 more per 1,000** (from 16 more to 96 more) | ⊕⊕⊕⊕ High | IMPORTANT |
| **Serious adverse events** | | | | | | | | | | | |
| 3 | RCTs | not serious | not serious | not serious | serious[b] | 73/755 (9.7%) | 74/750 (9.9%) | **RR 0.98** (0.72 to 1.33) | **2 fewer per 1,000** (from 28 fewer to 33 more) | ⊕⊕⊕◯ Moderate[b] | IMPORTANT |
| **Any gastrointestinal disorders** | | | | | | | | | | | |
| 3 | RCTs | not serious | not serious | not serious | not serious | 489/755 (64.8%) | 399/750 (53.2%) | **RR 1.22** (1.12 to 1.32) | **117 more per 1,000** (from 64 more to 170 more) | ⊕⊕⊕⊕ High | IMPORTANT |
| **Adverse events leading to permanent treatment discontinuation** | | | | | | | | | | | |
| 3 | RCTs | not serious | not serious | not serious | serious[b] | 59/755 (7.8%) | 55/750 (7.3%) | **RR 0.98** (0.37 to 2.61) | **1 fewer per 1,000** (from 46 fewer to 118 more) | ⊕⊕⊕◯ Moderate[b] | IMPORTANT |

**CI:** confidence interval; **MD:** mean difference; **MID:** minimally important difference; **RCTs:** randomized controlled trials; **RoM:** ratio of geometric means; **RR:** risk ratio.

**Explanations**

a. Downgraded one level: the 95% CI crossed the prespecified MID (0.3 percentage points).

b. Downgraded one level: the 95% CI crossed the null effect (no prespecified MID).

**Note to Figs. S1–S36** In all forest plots, squares represent study-specific estimates (size proportional to study weight), horizontal lines their 95% CIs, and diamonds the pooled estimates. Unless otherwise stated, pooled estimates were obtained with random-effects models, with HKSJ-adjusted CIs when $\tau^2 > 0$ and Wald-type CIs otherwise. All comparisons are CagriSema 2.4/2.4 mg versus CagriSema 1.0/1.0 mg. CI, confidence interval; HbA1c, glycated hemoglobin; HKSJ, Hartung–Knapp–Sidik–Jonkman; IV, inverse variance; MD, mean difference; MH, Mantel–Haenszel; OR, odds ratio; RoM, ratio of geometric means; RR, risk ratio; SD, standard deviation

| Studies | CagriSema 2.4/2.4 mg Mean | SD | Total | CagriSema 1.0/1.0 mg Mean | SD | Total | Weight | MD | 95% CI |
|---|---|---|---|---|---|---|---|---|---|
| Aroda 2026 | -13.6 | 7.09 | 62 | -11.5 | 7.14 | 63 | 11.2% | -2.10 | [-4.59; 0.39] |
| Buse 2026 | -12.9 | 8.87 | 603 | -11.2 | 8.41 | 595 | 72.8% | -1.70 | [-2.68; -0.72] |
| Rosenstock 2026 | -11.3 | 6.64 | 90 | -9.1 | 7.71 | 93 | 16.0% | -2.20 | [-4.28; -0.12] |
| **Total (95% CI)** | | | **755** | | | **751** | **100.0%** | **-1.83** | **[-2.66; -0.99]** |

Heterogeneity: $Tau^2 = 0$; $Chi^2 = 0.23$, df = 2 (P = 0.8897); $I^2 = 0.0\%$

Test for overall effect: Z = -4.29 (P < 0.001)

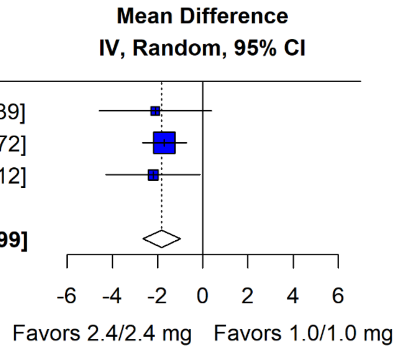


**Fig. S1** Percentage change in body weight, treatment-policy estimand (sensitivity analysis): CagriSema 2.4/2.4 mg was associated with a significantly greater reduction than CagriSema 1.0/1.0 mg. Pooled MD −1.83 percentage points (95% CI −2.66 to −0.99)

| Studies | CagriSema 2.4/2.4 mg Events | Total | CagriSema 1.0/1.0 mg Events | Total | Weight | RR | 95% CI |
|---|---|---|---|---|---|---|---|
| Aroda 2026 | 56 | 62 | 48 | 63 | 16.1% | 1.19 | [1.01; 1.39] |
| Buse 2026 | 522 | 603 | 497 | 595 | 68.6% | 1.04 | [0.99; 1.09] |
| Rosenstock 2026 | 70 | 90 | 68 | 93 | 15.3% | 1.06 | [0.90; 1.26] |
| **Total (95% CI)** | **648** | **755** | **613** | **751** | **100.0%** | **1.06** | **[0.92; 1.23]** |

Heterogeneity: $Tau^2 = 0.0013$; $Chi^2 = 2.51$, df = 2 (P = 0.2846); $I^2 = 24.9\%$

Test for overall effect: $t_2 = 1.79$ (P = 0.216)

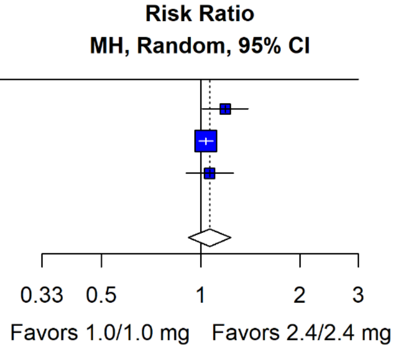


**Fig. S2** Proportion achieving ≥5% body-weight reduction: no statistically significant difference between doses. Pooled RR 1.06 (95% CI 0.92 to 1.23; HKSJ-adjusted). Sensitivity analysis with a Wald-type CI: 0.99 to 1.14

| Studies | CagriSema 2.4/2.4 mg Events | Total | CagriSema 1.0/1.0 mg Events | Total | Weight | RR | 95% CI |
|---|---|---|---|---|---|---|---|
| Aroda 2026 | 14 | 62 | 12 | 63 | 13.7% | 1.19 | [0.60; 2.36] |
| Buse 2026 | 143 | 603 | 89 | 595 | 78.3% | 1.59 | [1.25; 2.01] |
| Rosenstock 2026 | 8 | 90 | 9 | 93 | 8.0% | 0.92 | [0.37; 2.28] |
| **Total (95% CI)** | **165** | **755** | **110** | **751** | **100.0%** | **1.46** | **[0.87; 2.44]** |

Heterogeneity: $Tau^2 = 0.0080$; $Chi^2 = 1.78$, df = 2 (P = 0.4113); $I^2 = 8.2\%$
Test for overall effect: $t_2 = 3.16$ (P = 0.087)

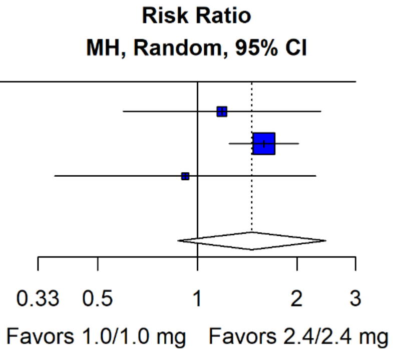


**Fig. S3** Proportion achieving ≥20% body-weight reduction: no statistically significant difference between doses. Pooled RR 1.46 (95% CI 0.87 to 2.44; HKSJ-adjusted). Sensitivity analysis with a Wald-type CI: 1.12 to 1.90

| Studies | CagriSema 2.4/2.4 mg Mean | SD | Total | CagriSema 1.0/1.0 mg Mean | SD | Total | Weight | MD | 95% CI |
|---|---|---|---|---|---|---|---|---|---|
| Aroda 2026 | -1.8 | 0.79 | 62 | -1.5 | 0.79 | 63 | 27.7% | -0.30 | [-0.58; -0.02] |
| Buse 2026 | -1.8 | 0.98 | 603 | -1.8 | 0.93 | 595 | 44.7% | -0.02 | [-0.13; 0.09] |
| Rosenstock 2026 | -2.3 | 0.85 | 90 | -1.9 | 1.06 | 93 | 27.6% | -0.33 | [-0.61; -0.05] |
| **Total (95% CI)** | | | **755** | | | **751** | **100.0%** | **-0.18** | **[-0.63; 0.26]** |

Heterogeneity: $Tau^2 = 0.0246$; $Chi^2 = 6.67$, df = 2 (P = 0.0356); $I^2 = 67.0\%$
Test for overall effect: $t_2 = -1.76$ (P = 0.220)

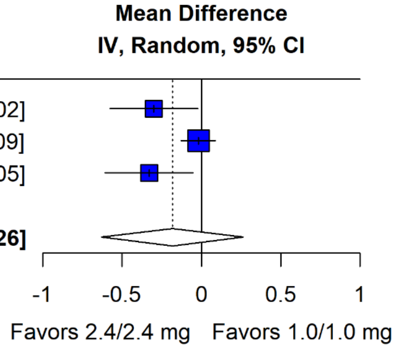


**Fig. S4** Change in HbA1c (percentage points), treatment-policy estimand (sensitivity analysis): no statistically significant difference between doses. Pooled MD −0.18 percentage points [−2.0 mmol/mol] (95% CI −0.63 to 0.26 [−6.9 to 2.8 mmol/mol]; HKSJ-adjusted). Sensitivity analysis with a Wald-type CI: −0.40 to 0.03 (−4.4 to 0.3 mmol/mol)

| Studies | CagriSema 2.4/2.4 mg Events | Total | CagriSema 1.0/1.0 mg Events | Total | Weight | RR | 95% CI |
|---|---|---|---|---|---|---|---|
| Aroda 2026 | 50 | 62 | 47 | 63 | 12.4% | 1.08 | [0.90; 1.31] |
| Buse 2026 | 437 | 603 | 408 | 595 | 82.0% | 1.06 | [0.98; 1.14] |
| Rosenstock 2026 | 52 | 90 | 43 | 93 | 5.6% | 1.25 | [0.94; 1.66] |
| **Total (95% CI)** | **539** | **755** | **498** | **751** | **100.0%** | **1.07** | **[1.00; 1.14]** |

Heterogeneity: $Tau^2 = 0$; $Chi^2 = 1.29$, df = 2 (P = 0.5253); $I^2 = 0.0\%$
Test for overall effect: Z = 1.99 (P = 0.047)

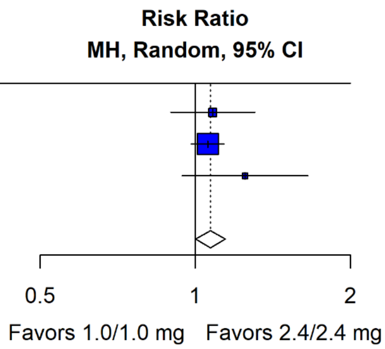


**Fig. S5** Proportion achieving HbA1c ≤6.5% (≤48 mmol/mol): CagriSema 2.4/2.4 mg was associated with a significantly higher proportion than CagriSema 1.0/1.0 mg. Pooled RR 1.07 (95% CI 1.00 to 1.14)

| Studies | CagriSema 2.4/2.4 mg Events | Total | CagriSema 1.0/1.0 mg Events | Total | Weight | RR | 95% CI |
|---|---|---|---|---|---|---|---|
| Aroda 2026 | 54 | 62 | 51 | 63 | 10.2% | 1.08 | [0.92; 1.25] |
| Buse 2026 | 496 | 603 | 481 | 595 | 82.3% | 1.02 | [0.96; 1.07] |
| Rosenstock 2026 | 68 | 90 | 65 | 93 | 7.6% | 1.08 | [0.90; 1.29] |
| **Total (95% CI)** | **618** | **755** | **597** | **751** | **100.0%** | **1.03** | **[0.98; 1.08]** |

Heterogeneity: $Tau^2 = 0$; $Chi^2 = 0.79$, df = 2 (P = 0.6752); $I^2 = 0.0\%$
Test for overall effect: Z = 1.11 (P = 0.268)

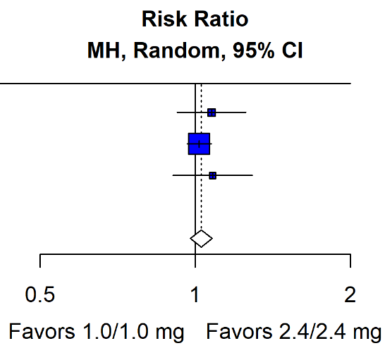


**Fig. S6** Proportion achieving HbA1c <7.0% (<53 mmol/mol): no statistically significant difference between doses. Pooled RR 1.03 (95% CI 0.98 to 1.08)

| Studies | CagriSema 2.4/2.4 mg Events | Total | CagriSema 1.0/1.0 mg Events | Total | Weight | RR | 95% CI |
|---|---|---|---|---|---|---|---|
| Aroda 2026 | 0 | 62 | 3 | 63 | 15.7% | 0.15 | [0.01; 2.75] |
| Buse 2026 | 27 | 603 | 37 | 595 | 51.7% | 0.72 | [0.44; 1.17] |
| Rosenstock 2026 | 7 | 90 | 2 | 93 | 32.6% | 3.62 | [0.77; 16.95] |
| **Total (95% CI)** | **34** | **755** | **42** | **751** | **100.0%** | **0.95** | **[0.03; 25.70]** |

Heterogeneity: $Tau^2 = 0.8958$; $Chi^2 = 5.14$, df = 2 (P = 0.0765); $I^2 = 62.6\%$
Test for overall effect: $t_2 = -0.07$ (P = 0.951)

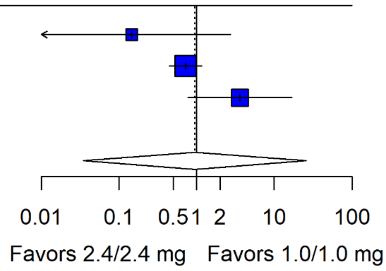


**Fig. S7** Initiation of glycemic rescue medication: no statistically significant difference between doses. Pooled RR 0.95 (95% CI 0.03 to 25.70; HKSJ-adjusted). Sensitivity analysis with a Wald-type CI: 0.24 to 3.76. In REIMAGINE 1, rescue medication was reported only in the text (two participants initiated metformin and one a sulfonylurea in the 1.0/1.0 mg arm); these were summed for analysis (three participants)

| Studies | CagriSema 2.4/2.4 mg Mean | SD | Total | CagriSema 1.0/1.0 mg Mean | SD | Total | Weight | MD | 95% CI |
|---|---|---|---|---|---|---|---|---|---|
| Aroda 2026 | -3.7 | 7.09 | 62 | -2.5 | 7.14 | 63 | 11.3% | -1.20 | [-3.69; 1.29] |
| Buse 2026 | -3.2 | 8.42 | 603 | -2.7 | 8.41 | 595 | 77.5% | -0.50 | [-1.45; 0.45] |
| Rosenstock 2026 | -1.5 | 9.49 | 90 | -2.5 | 7.71 | 93 | 11.2% | 1.00 | [-1.51; 3.51] |
| **Total (95% CI)** | | | **755** | | | **751** | **100.0%** | **-0.41** | **[-1.25; 0.43]** |

Heterogeneity: $Tau^2 < 0.0001$; $Chi^2 = 1.63$, df = 2 (P = 0.4423); $I^2 = 0.0\%$
Test for overall effect: Z = -0.96 (P = 0.336)

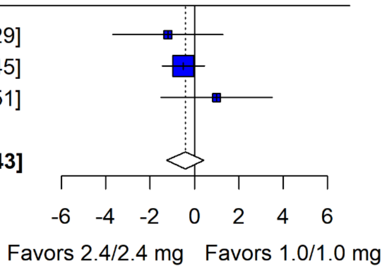


**Fig. S8** Change in diastolic blood pressure (mmHg): no statistically significant difference between doses. Pooled MD −0.41 mmHg (95% CI −1.25 to 0.43)

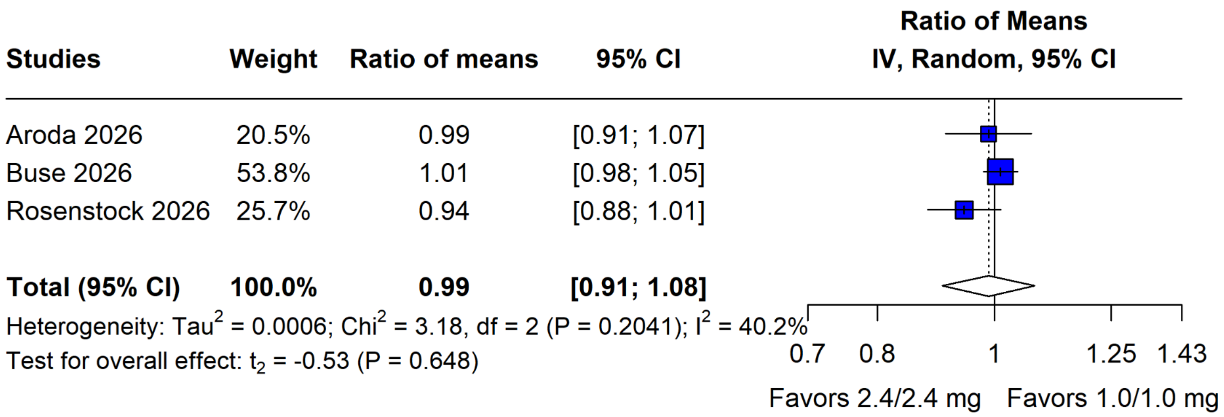


**Fig. S9** Non-HDL cholesterol (ratio to baseline): no statistically significant difference between doses. Pooled RoM 0.99 (95% CI 0.91 to 1.08; HKSJ-adjusted). Sensitivity analysis with a Wald-type CI: 0.95 to 1.03. HDL, high-density lipoprotein

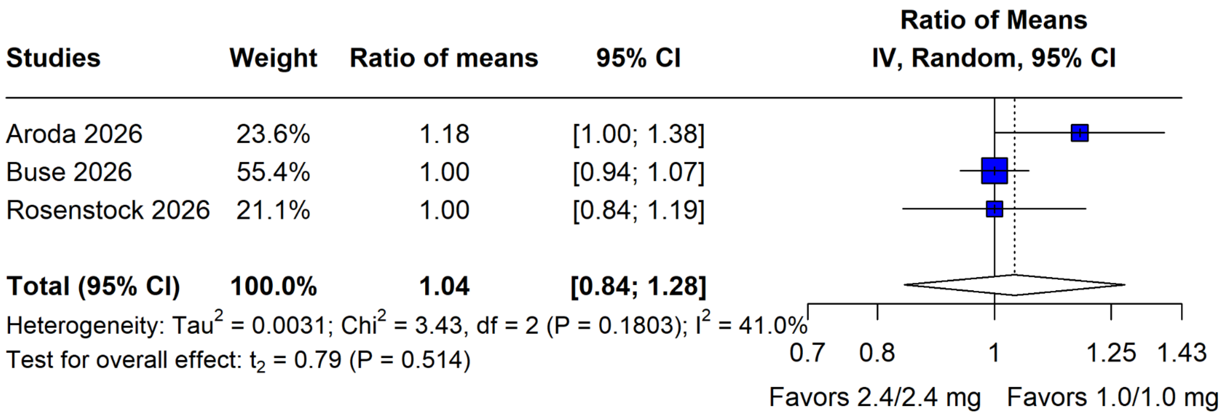


**Fig. S10** Free fatty acids (ratio to baseline): no statistically significant difference between doses. Pooled RoM 1.04 (95% CI 0.84 to 1.28; HKSJ-adjusted). Sensitivity analysis with a Wald-type CI: 0.95 to 1.14

| Studies | Weight | Ratio of means | 95% CI |
|---|---|---|---|
| Aroda 2026 | 9.9% | 1.04 | [0.98; 1.10] |
| Buse 2026 | 80.2% | 1.02 | [1.00; 1.04] |
| Rosenstock 2026 | 9.9% | 1.02 | [0.96; 1.08] |
| **Total (95% CI)** | **100.0%** | **1.02** | **[1.00; 1.04]** |

Heterogeneity: $Tau^2$ = 0; $Chi^2$ = 0.43, df = 2 (P = 0.8050); $I^2$ = 0.0%
Test for overall effect: Z = 2.26 (P = 0.024)

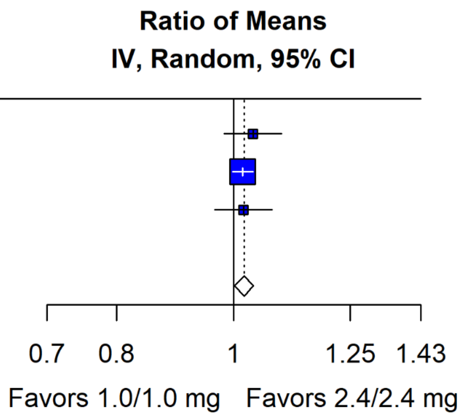


**Fig. S11** HDL cholesterol (ratio to baseline): CagriSema 2.4/2.4 mg was associated with a significantly greater increase than CagriSema 1.0/1.0 mg. Pooled RoM 1.02 (95% CI 1.00 to 1.04). HDL, high-density lipoprotein

| Studies | Weight | Ratio of means | 95% CI |
|---|---|---|---|
| Aroda 2026 | 8.5% | 0.99 | [0.86; 1.13] |
| Buse 2026 | 74.8% | 0.96 | [0.92; 1.00] |
| Rosenstock 2026 | 16.7% | 0.92 | [0.84; 1.02] |
| **Total (95% CI)** | **100.0%** | **0.96** | **[0.92; 0.99]** |

Heterogeneity: $Tau^2$ = 0; $Chi^2$ = 0.68, df = 2 (P = 0.7125); $I^2$ = 0.0%
Test for overall effect: Z = -2.29 (P = 0.022)

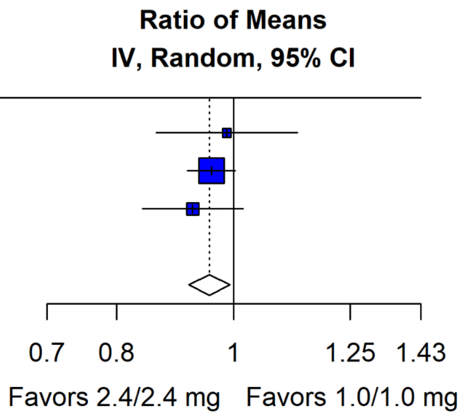


**Fig. S12** Triglycerides (ratio to baseline): CagriSema 2.4/2.4 mg was associated with a significantly greater reduction than CagriSema 1.0/1.0 mg. Pooled RoM 0.96 (95% CI 0.92 to 0.99)

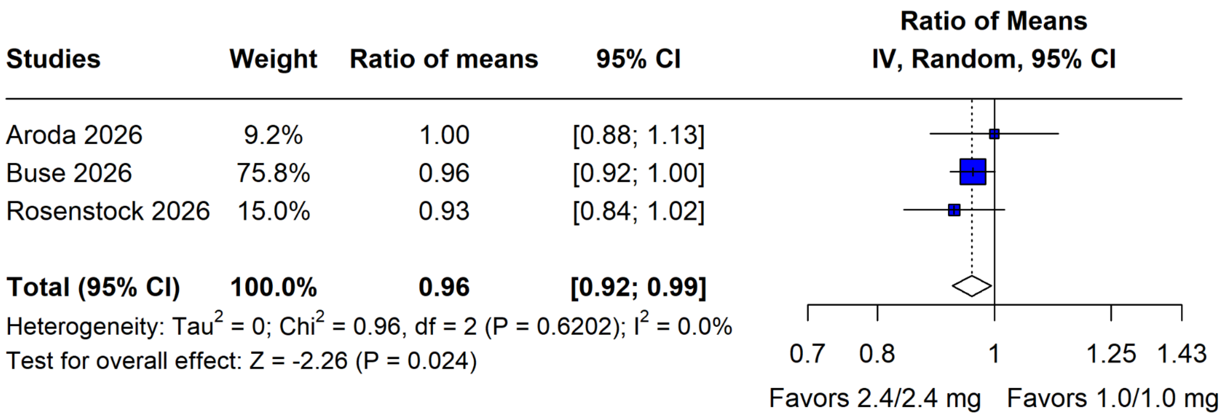


**Fig. S13** VLDL cholesterol (ratio to baseline): CagriSema 2.4/2.4 mg was associated with a significantly greater reduction than CagriSema 1.0/1.0 mg. Pooled RoM 0.96 (95% CI 0.92 to 0.99). VLDL, very-low-density lipoprotein

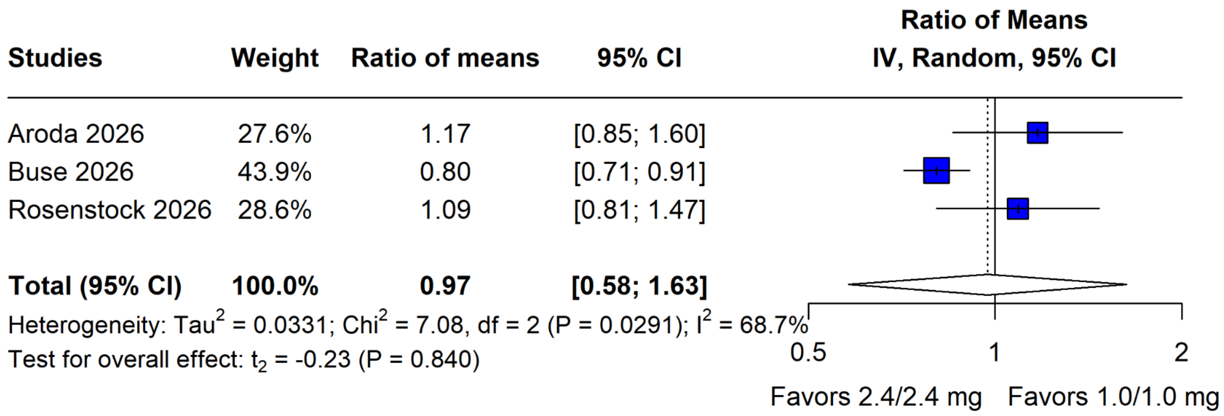


**Fig. S14** High-sensitivity C-reactive protein (ratio to baseline): no statistically significant difference between doses. Pooled RoM 0.97 (95% CI 0.58 to 1.63; HKSJ-adjusted). Sensitivity analysis with a Wald-type CI: 0.76 to 1.25

| Studies | CagriSema 2.4/2.4 mg Mean | SD | Total | CagriSema 1.0/1.0 mg Mean | SD | Total | Weight | MD | 95% CI |
|---|---|---|---|---|---|---|---|---|---|
| Aroda 2026 | 2.0 | 7.09 | 62 | 2.8 | 7.94 | 63 | 7.9% | -0.80 | [-3.44; 1.84] |
| Buse 2026 | 2.5 | 7.54 | 603 | 2.3 | 7.52 | 595 | 75.3% | 0.20 | [-0.65; 1.05] |
| Rosenstock 2026 | 1.4 | 6.64 | 90 | 1.7 | 5.79 | 93 | 16.8% | -0.30 | [-2.11; 1.51] |
| **Total (95% CI)** | | | **755** | | | **751** | **100.0%** | **0.04** | **[-0.70; 0.78]** |

Heterogeneity: $Tau^2 = 0$; $Chi^2 = 0.66$, df = 2 (P = 0.7186); $I^2 = 0.0\%$
Test for overall effect: Z = 0.10 (P = 0.921)

**Mean Difference IV, Random, 95% CI**

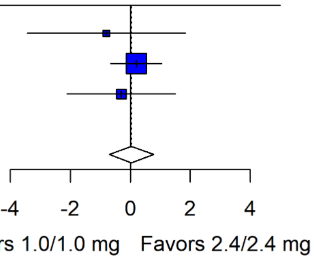


**Fig. S15** Change in SF-36v2 physical component summary score: no statistically significant difference between doses. Pooled MD 0.04 (95% CI −0.70 to 0.78). SF-36v2, 36-Item Short Form Health Survey version 2

| Studies | CagriSema 2.4/2.4 mg Mean | SD | Total | CagriSema 1.0/1.0 mg Mean | SD | Total | Weight | MD | 95% CI |
|---|---|---|---|---|---|---|---|---|---|
| Aroda 2026 | -0.5 | 8.66 | 62 | -0.4 | 9.52 | 63 | 18.3% | -0.10 | [-3.29; 3.09] |
| Buse 2026 | 0.8 | 9.31 | 603 | 0.0 | 9.29 | 595 | 54.2% | 0.80 | [-0.25; 1.85] |
| Rosenstock 2026 | -0.5 | 8.54 | 90 | 1.1 | 7.71 | 93 | 27.5% | -1.60 | [-3.96; 0.76] |
| **Total (95% CI)** | | | **755** | | | **751** | **100.0%** | **-0.03** | **[-3.15; 3.10]** |

Heterogeneity: $Tau^2 = 0.9114$; $Chi^2 = 3.40$, df = 2 (P = 0.1827); $I^2 = 44.3\%$
Test for overall effect: $t_2 = -0.04$ (P = 0.975)

**Mean Difference IV, Random, 95% CI**

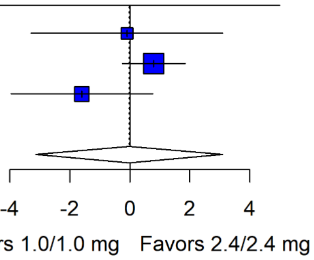


**Fig. S16** Change in SF-36v2 mental component summary score: no statistically significant difference between doses. Pooled MD −0.03 (95% CI −3.15 to 3.10; HKSJ-adjusted). Sensitivity analysis with a Wald-type CI: −1.61 to 1.56. SF-36v2, 36-Item Short Form Health Survey version 2

| Studies | CagriSema 2.4/2.4 mg Mean | SD | Total | CagriSema 1.0/1.0 mg Mean | SD | Total | Weight | MD | 95% CI |
|---|---|---|---|---|---|---|---|---|---|
| Aroda 2026 | 7.7 | 5.51 | 62 | 8.3 | 5.56 | 63 | 10.4% | -0.60 | [-2.54; 1.34] |
| Buse 2026 | 5.5 | 6.21 | 603 | 5.4 | 6.20 | 595 | 79.2% | 0.10 | [-0.60; 0.80] |
| Rosenstock 2026 | 4.3 | 6.64 | 90 | 5.3 | 6.75 | 93 | 10.4% | -1.00 | [-2.94; 0.94] |
| **Total (95% CI)** | | | **755** | | | **751** | **100.0%** | **-0.09** | **[-0.71; 0.54]** |

Heterogeneity: $Tau^2 = 0$; $Chi^2 = 1.39$, df = 2 (P = 0.4987); $I^2 = 0.0\%$
Test for overall effect: Z = -0.27 (P = 0.785)

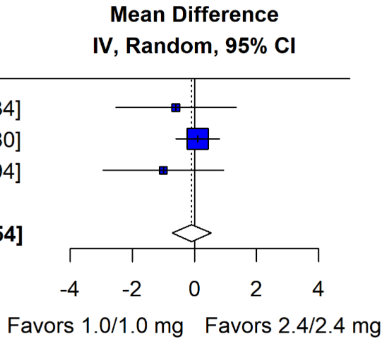


**Fig. S17** Change in DTSQ score: no statistically significant difference between doses. Pooled MD −0.09 (95% CI −0.71 to 0.54). DTSQ, Diabetes Treatment Satisfaction Questionnaire

| Studies | CagriSema 2.4/2.4 mg Events | Total | CagriSema 1.0/1.0 mg Events | Total | Weight | RR | 95% CI |
|---|---|---|---|---|---|---|---|
| Aroda 2026 | 1 | 62 | 1 | 63 | 15.1% | 1.02 | [0.06; 15.89] |
| Buse 2026 | 27 | 603 | 18 | 594 | 53.4% | 1.48 | [0.82; 2.65] |
| Rosenstock 2026 | 2 | 90 | 8 | 93 | 31.5% | 0.26 | [0.06; 1.18] |
| **Total (95% CI)** | **30** | **755** | **27** | **750** | **100.0%** | **0.81** | **[0.07; 8.71]** |

Heterogeneity: $Tau^2 = 0.6519$; $Chi^2 = 4.40$, df = 2 (P = 0.1106); $I^2 = 55.0\%$
Test for overall effect: $t_2 = -0.39$ (P = 0.734)

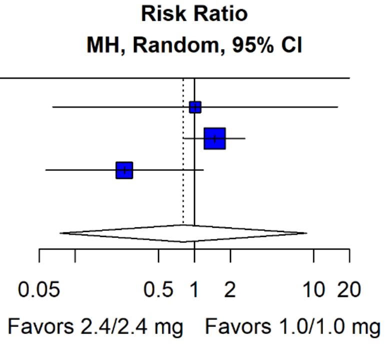


**Fig. S18** Gastrointestinal adverse events leading to permanent treatment discontinuation: no statistically significant difference between doses. Pooled RR 0.81 (95% CI 0.07 to 8.71; HKSJ-adjusted). Sensitivity analysis with a Wald-type CI: 0.23 to 2.76

| Studies | CagriSema 2.4/2.4 mg Events | Total | CagriSema 1.0/1.0 mg Events | Total | Weight | RR | 95% CI |
|---|---|---|---|---|---|---|---|
| Aroda 2026 | 22 | 62 | 17 | 63 | 8.0% | 1.31 | [0.78; 2.23] |
| Buse 2026 | 223 | 603 | 168 | 594 | 81.3% | 1.31 | [1.11; 1.54] |
| Rosenstock 2026 | 28 | 90 | 25 | 93 | 10.7% | 1.16 | [0.73; 1.82] |
| **Total (95% CI)** | **273** | **755** | **210** | **750** | **100.0%** | **1.29** | **[1.11; 1.50]** |

Heterogeneity: $Tau^2 = 0$; $Chi^2 = 0.25$, df = 2 (P = 0.8827); $I^2 = 0.0\%$
Test for overall effect: Z = 3.36 (P < 0.001)

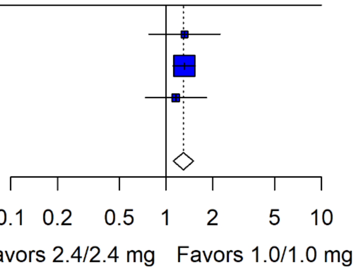


**Fig. S19** Nausea: CagriSema 2.4/2.4 mg was associated with a significantly higher incidence than CagriSema 1.0/1.0 mg. Pooled RR 1.29 (95% CI 1.11 to 1.50)

| Studies | CagriSema 2.4/2.4 mg Events | Total | CagriSema 1.0/1.0 mg Events | Total | Weight | RR | 95% CI |
|---|---|---|---|---|---|---|---|
| Aroda 2026 | 9 | 62 | 5 | 63 | 6.2% | 1.83 | [0.65; 5.15] |
| Buse 2026 | 96 | 603 | 73 | 594 | 83.4% | 1.30 | [0.98; 1.72] |
| Rosenstock 2026 | 13 | 90 | 9 | 93 | 10.4% | 1.49 | [0.67; 3.32] |
| **Total (95% CI)** | **118** | **755** | **87** | **750** | **100.0%** | **1.34** | **[1.04; 1.74]** |

Heterogeneity: $Tau^2 = 0$; $Chi^2 = 0.47$, df = 2 (P = 0.7900); $I^2 = 0.0\%$
Test for overall effect: Z = 2.24 (P = 0.025)

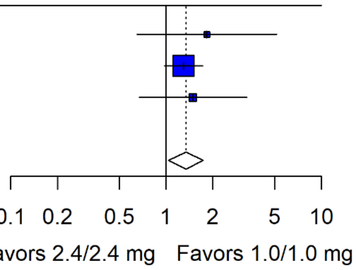


**Fig. S20** Vomiting: CagriSema 2.4/2.4 mg was associated with a significantly higher incidence than CagriSema 1.0/1.0 mg. Pooled RR 1.34 (95% CI 1.04 to 1.74)

| Studies | CagriSema 2.4/2.4 mg Events | Total | CagriSema 1.0/1.0 mg Events | Total | Weight | RR | 95% CI |
|---|---|---|---|---|---|---|---|
| Aroda 2026 | 17 | 62 | 6 | 63 | 8.6% | 2.88 | [1.22; 6.82] |
| Buse 2026 | 95 | 603 | 67 | 594 | 75.5% | 1.40 | [1.04; 1.87] |
| Rosenstock 2026 | 18 | 90 | 14 | 93 | 15.9% | 1.33 | [0.70; 2.51] |
| **Total (95% CI)** | **130** | **755** | **87** | **750** | **100.0%** | **1.47** | **[1.14; 1.90]** |

Heterogeneity: $Tau^2 < 0.0001$; $Chi^2 = 2.55$, df = 2 (P = 0.2794); $I^2 = 0.0\%$
Test for overall effect: Z = 3.01 (P = 0.003)

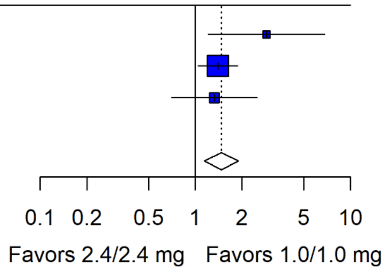


**Fig. S21** Decreased appetite: CagriSema 2.4/2.4 mg was associated with a significantly higher incidence than CagriSema 1.0/1.0 mg. Pooled RR 1.47 (95% CI 1.14 to 1.90)

| Studies | CagriSema 2.4/2.4 mg Events | Total | CagriSema 1.0/1.0 mg Events | Total | Weight | RR | 95% CI |
|---|---|---|---|---|---|---|---|
| Aroda 2026 | 7 | 62 | 4 | 63 | 8.9% | 1.78 | [0.55; 5.77] |
| Buse 2026 | 61 | 603 | 40 | 594 | 84.1% | 1.50 | [1.02; 2.20] |
| Rosenstock 2026 | 7 | 90 | 3 | 93 | 7.0% | 2.41 | [0.64; 9.04] |
| **Total (95% CI)** | **75** | **755** | **47** | **750** | **100.0%** | **1.58** | **[1.11; 2.24]** |

Heterogeneity: $Tau^2 = 0$; $Chi^2 = 0.50$, df = 2 (P = 0.7793); $I^2 = 0.0\%$
Test for overall effect: Z = 2.54 (P = 0.011)

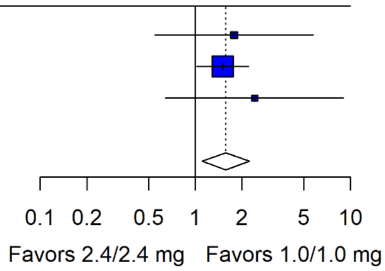


**Fig. S22** Fatigue: CagriSema 2.4/2.4 mg was associated with a significantly higher incidence than CagriSema 1.0/1.0 mg. Pooled RR 1.58 (95% CI 1.11 to 2.24)

| Studies | CagriSema 2.4/2.4 mg Events | Total | CagriSema 1.0/1.0 mg Events | Total | Weight | RR | 95% CI |
|---|---|---|---|---|---|---|---|
| Aroda 2026 | 9 | 62 | 8 | 63 | 5.3% | 1.14 | [0.47; 2.77] |
| Buse 2026 | 136 | 603 | 119 | 594 | 87.4% | 1.13 | [0.90; 1.40] |
| Rosenstock 2026 | 14 | 90 | 10 | 93 | 7.3% | 1.45 | [0.68; 3.09] |
| **Total (95% CI)** | **159** | **755** | **137** | **750** | **100.0%** | **1.15** | **[0.94; 1.41]** |

Heterogeneity: $Tau^2 = 0$; $Chi^2 = 0.39$, df = 2 (P = 0.8235); $I^2 = 0.0\%$
Test for overall effect: Z = 1.32 (P = 0.187)

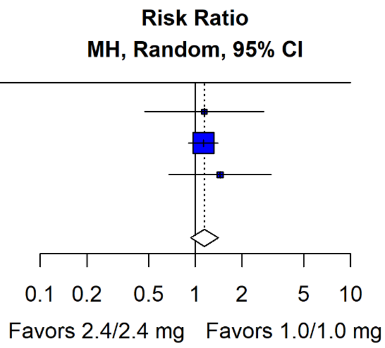


**Fig. S23** Diarrhea: no statistically significant difference between doses. Pooled RR 1.15 (95% CI 0.94 to 1.41)

| Studies | CagriSema 2.4/2.4 mg Events | Total | CagriSema 1.0/1.0 mg Events | Total | Weight | RR | 95% CI |
|---|---|---|---|---|---|---|---|
| Aroda 2026 | 9 | 62 | 11 | 63 | 7.4% | 0.83 | [0.37; 1.87] |
| Buse 2026 | 126 | 603 | 99 | 594 | 85.2% | 1.25 | [0.99; 1.59] |
| Rosenstock 2026 | 10 | 90 | 11 | 93 | 7.4% | 0.94 | [0.42; 2.10] |
| **Total (95% CI)** | **145** | **755** | **121** | **750** | **100.0%** | **1.19** | **[0.96; 1.48]** |

Heterogeneity: $Tau^2 = 0$; $Chi^2 = 1.27$, df = 2 (P = 0.5293); $I^2 = 0.0\%$
Test for overall effect: Z = 1.56 (P = 0.119)

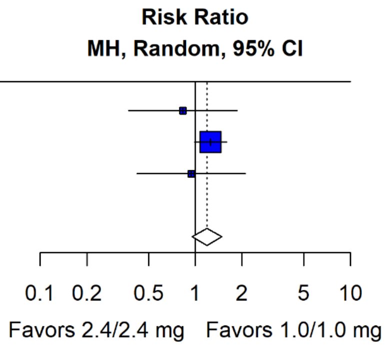


**Fig. S24** Constipation: no statistically significant difference between doses. Pooled RR 1.19 (95% CI 0.96 to 1.48)

| Studies | CagriSema 2.4/2.4 mg Events | Total | CagriSema 1.0/1.0 mg Events | Total | Weight | RR | 95% CI |
|---|---|---|---|---|---|---|---|
| Aroda 2026 | 2 | 62 | 2 | 63 | 10.1% | 1.02 | [0.15; 6.99] |
| Buse 2026 | 25 | 603 | 13 | 594 | 86.2% | 1.89 | [0.98; 3.67] |
| Rosenstock 2026 | 1 | 90 | 0 | 93 | 3.7% | 3.10 | [0.13; 75.10] |
| **Total (95% CI)** | **28** | **755** | **15** | **750** | **100.0%** | **1.81** | **[0.98; 3.34]** |

Heterogeneity: $Tau^2 = 0$; $Chi^2 = 0.47$, df = 2 (P = 0.7898); $I^2 = 0.0\%$
Test for overall effect: Z = 1.90 (P = 0.058)

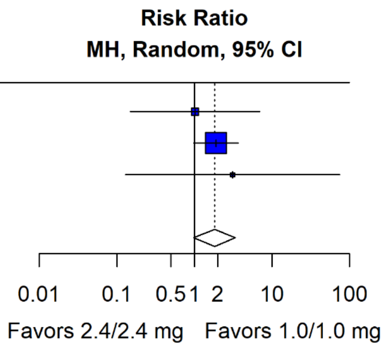


**Fig. S25** Injection-site reactions: no statistically significant difference between doses. Pooled RR 1.81 (95% CI 0.98 to 3.34)

| Studies | CagriSema 2.4/2.4 mg Events | Total | CagriSema 1.0/1.0 mg Events | Total | Weight | RR | 95% CI |
|---|---|---|---|---|---|---|---|
| Aroda 2026 | 0 | 62 | 3 | 63 | 0.6% | 0.15 | [0.01; 2.75] |
| Buse 2026 | 64 | 603 | 66 | 594 | 53.3% | 0.96 | [0.69; 1.32] |
| Rosenstock 2026 | 40 | 90 | 35 | 93 | 46.1% | 1.18 | [0.83; 1.67] |
| **Total (95% CI)** | **104** | **755** | **104** | **750** | **100.0%** | **1.04** | **[0.82; 1.32]** |

Heterogeneity: $Tau^2 < 0.0001$; $Chi^2 = 2.49$, df = 2 (P = 0.2874); $I^2 = 0.0\%$
Test for overall effect: Z = 0.33 (P = 0.742)

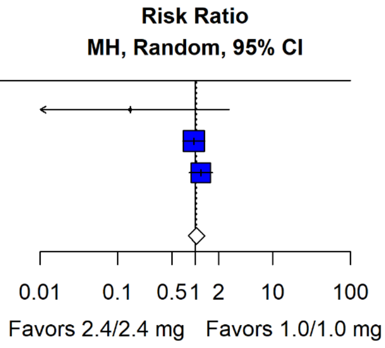


**Fig. S26** Level 1 hypoglycemia: no statistically significant difference between doses. Pooled RR 1.04 (95% CI 0.82 to 1.32)

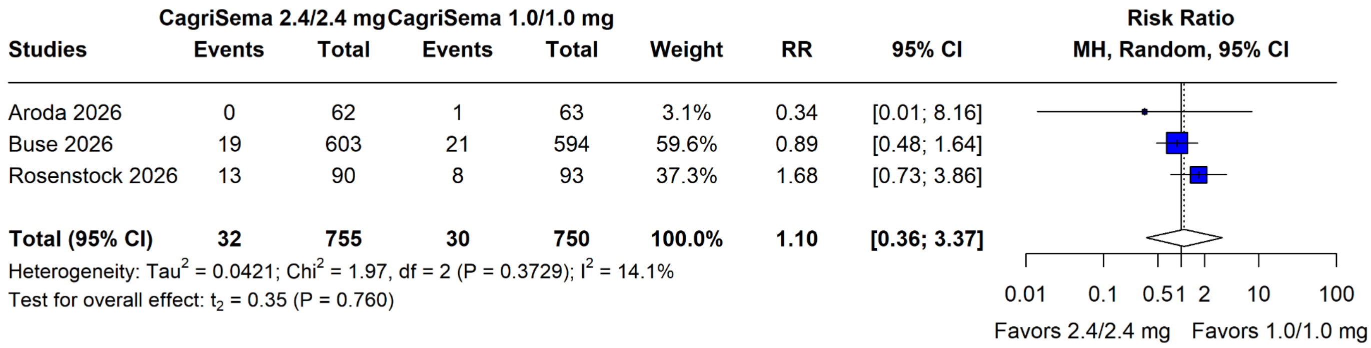


**Fig. S27** Level 2 hypoglycemia: no statistically significant difference between doses. Pooled RR 1.10 (95% CI 0.36 to 3.37; HKSJ-adjusted). Sensitivity analysis with a Wald-type CI: 0.62 to 1.93

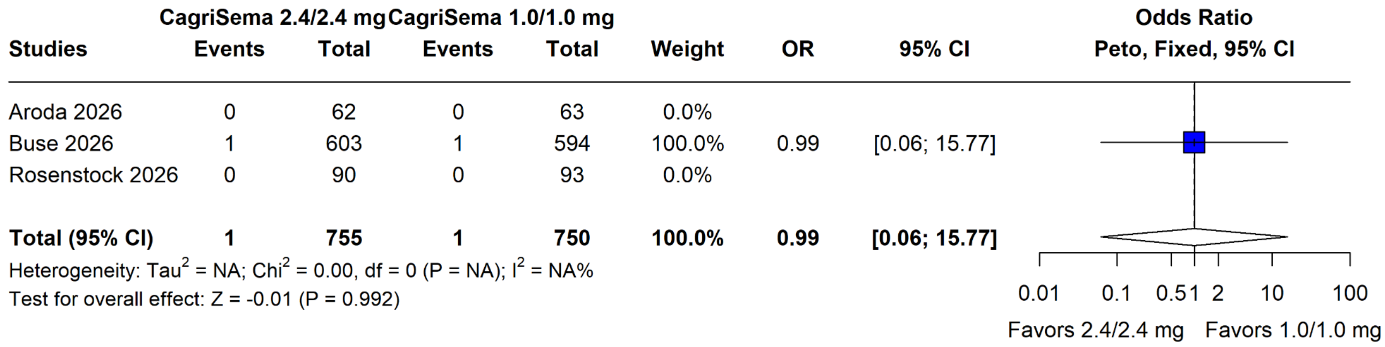


**Fig. S28** Level 3 hypoglycemia: no statistically significant difference between doses. Peto OR 0.99 (95% CI 0.06 to 15.77; common-effect model)

| Studies | CagriSema 2.4/2.4 mg Events | Total | CagriSema 1.0/1.0 mg Events | Total | Weight | RR | 95% CI |
|---|---|---|---|---|---|---|---|
| Aroda 2026 | 4 | 62 | 2 | 63 | 14.1% | 2.03 | [0.39; 10.70] |
| Buse 2026 | 50 | 603 | 47 | 594 | 57.0% | 1.05 | [0.72; 1.54] |
| Rosenstock 2026 | 13 | 90 | 5 | 93 | 28.9% | 2.69 | [1.00; 7.23] |
| **Total (95% CI)** | **67** | **755** | **54** | **750** | **100.0%** | **1.51** | **[0.41; 5.58]** |

Heterogeneity: $Tau^2 = 0.1857$; $Chi^2 = 3.41$, df = 2 (P = 0.1814); $I^2 = 46.0\%$
Test for overall effect: $t_2 = 1.36$ (P = 0.307)

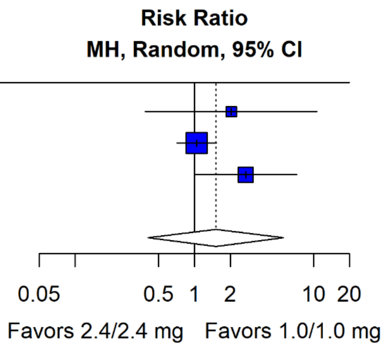


**Fig. S29** Retinal disorders: no statistically significant difference between doses. Pooled RR 1.51 (95% CI 0.41 to 5.58; HKSJ-adjusted). Sensitivity analysis with a Wald-type CI: 0.75 to 3.04

| Studies | CagriSema 2.4/2.4 mg Events | Total | CagriSema 1.0/1.0 mg Events | Total | Weight | RR | 95% CI |
|---|---|---|---|---|---|---|---|
| Aroda 2026 | 1 | 62 | 0 | 63 | 10.6% | 3.05 | [0.13; 73.40] |
| Buse 2026 | 14 | 603 | 11 | 594 | 76.8% | 1.25 | [0.57; 2.74] |
| Rosenstock 2026 | 4 | 90 | 0 | 93 | 12.5% | 9.30 | [0.51; 170.25] |
| **Total (95% CI)** | **19** | **755** | **11** | **750** | **100.0%** | **1.77** | **[0.22; 14.19]** |

Heterogeneity: $Tau^2 = 0.2392$; $Chi^2 = 1.91$, df = 2 (P = 0.3857); $I^2 = 15.4\%$
Test for overall effect: $t_2 = 1.18$ (P = 0.359)

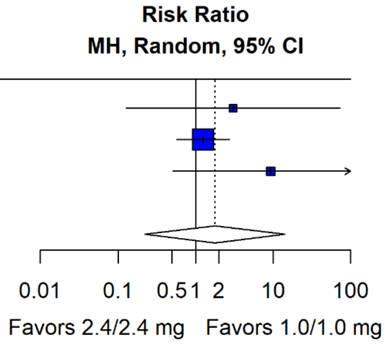


**Fig. S30** Gallbladder-related disorders: no statistically significant difference between doses. Pooled RR 1.77 (95% CI 0.22 to 14.19; HKSJ-adjusted). Sensitivity analysis with a Wald-type CI: 0.60 to 5.24

| Studies | CagriSema 2.4/2.4 mg Events | Total | CagriSema 1.0/1.0 mg Events | Total | Weight | RR | 95% CI |
|---|---|---|---|---|---|---|---|
| Aroda 2026 | 6 | 62 | 2 | 63 | 8.5% | 3.05 | [0.64; 14.53] |
| Buse 2026 | 33 | 603 | 37 | 594 | 73.4% | 0.88 | [0.56; 1.39] |
| Rosenstock 2026 | 7 | 90 | 6 | 93 | 18.1% | 1.21 | [0.42; 3.45] |
| **Total (95% CI)** | **46** | **755** | **45** | **750** | **100.0%** | **1.03** | **[0.36; 3.01]** |

Heterogeneity: $Tau^2 = 0.0225$; $Chi^2 = 2.39$, df = 2 (P = 0.3028); $I^2 = 8.6\%$
Test for overall effect: $t_2 = 0.14$ (P = 0.904)

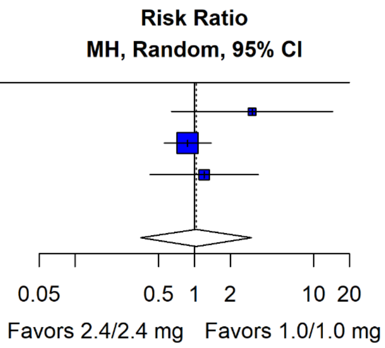


**Fig. S31** Neoplasms: no statistically significant difference between doses. Pooled RR 1.03 (95% CI 0.36 to 3.01; HKSJ-adjusted). Sensitivity analysis with a Wald-type CI: 0.65 to 1.65

| Studies | CagriSema 2.4/2.4 mg Events | Total | CagriSema 1.0/1.0 mg Events | Total | Weight | RR | 95% CI |
|---|---|---|---|---|---|---|---|
| Aroda 2026 | 0 | 62 | 0 | 63 | 0.0% | | |
| Buse 2026 | 9 | 603 | 6 | 594 | 84.3% | 1.48 | [0.53; 4.13] |
| Rosenstock 2026 | 1 | 90 | 2 | 93 | 15.7% | 0.52 | [0.05; 5.60] |
| **Total (95% CI)** | **10** | **755** | **8** | **750** | **100.0%** | **1.25** | **[0.49; 3.22]** |

Heterogeneity: $Tau^2 = 0$; $Chi^2 = 0.63$, df = 1 (P = 0.4273); $I^2 = 0.0\%$
Test for overall effect: Z = 0.47 (P = 0.639)

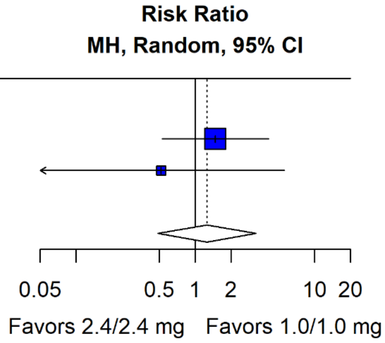


**Fig. S32** Malignant neoplasms: no statistically significant difference between doses. Pooled RR 1.25 (95% CI 0.49 to 3.22); Aroda 2026 (REIMAGINE 1) reported no events in either arm and did not contribute to the pooled estimate

| Studies | CagriSema 2.4/2.4 mg Events | Total | CagriSema 1.0/1.0 mg Events | Total | Weight | RR | 95% CI | Risk Ratio MH, Random, 95% CI |
|---|---|---|---|---|---|---|---|---|
| Aroda 2026 | 2 | 62 | 0 | 63 | 3.1% | 5.08 | [0.25; 103.70] | |
| Buse 2026 | 21 | 603 | 23 | 594 | 83.8% | 0.90 | [0.50; 1.61] | |
| Rosenstock 2026 | 4 | 90 | 3 | 93 | 13.1% | 1.38 | [0.32; 5.98] | |
| **Total (95% CI)** | **27** | **755** | **26** | **750** | **100.0%** | **1.00** | **[0.59; 1.71]** | |

Heterogeneity: $Tau^2 = 0$; $Chi^2 = 1.43$, df = 2 (P = 0.4901); $I^2 = 0.0\%$
Test for overall effect: Z = 0.01 (P = 0.989)

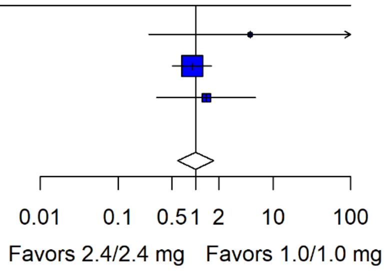


**Fig. S33** Hypersensitivity reactions: no statistically significant difference between doses. Pooled RR 1.00 (95% CI 0.59 to 1.71)

| Studies | CagriSema 2.4/2.4 mg Events | Total | CagriSema 1.0/1.0 mg Events | Total | Weight | OR | 95% CI | Odds Ratio Peto, Fixed, 95% CI |
|---|---|---|---|---|---|---|---|---|
| Aroda 2026 | 0 | 62 | 0 | 63 | 0.0% | | | |
| Buse 2026 | 4 | 603 | 2 | 594 | 85.7% | 1.92 | [0.39; 9.56] | |
| Rosenstock 2026 | 0 | 90 | 1 | 93 | 14.3% | 0.14 | [0.00; 7.05] | |
| **Total (95% CI)** | **4** | **755** | **3** | **750** | **100.0%** | **1.32** | **[0.30; 5.83]** | |

Heterogeneity: $Tau^2 = 1.1025$; $Chi^2 = 1.47$, df = 1 (P = 0.2250); $I^2 = 32.1\%$
Test for overall effect: Z = 0.37 (P = 0.713)

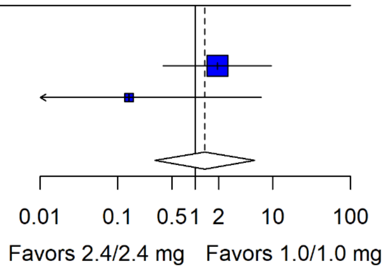


**Fig. S34** Deaths: no statistically significant difference between doses. Peto OR 1.32 (95% CI 0.30 to 5.83; common-effect model); Aroda 2026 (REIMAGINE 1) reported no events in either arm

| Studies | CagriSema 2.4/2.4 mg Events | Total | CagriSema 1.0/1.0 mg Events | Total | Weight | RR | 95% CI |
|---|---|---|---|---|---|---|---|
| Aroda 2026 | 47 | 62 | 59 | 63 | 27.1% | 0.81 | [0.69; 0.94] |
| Buse 2026 | 437 | 603 | 555 | 595 | 43.0% | 0.78 | [0.74; 0.82] |
| Rosenstock 2026 | 72 | 90 | 78 | 93 | 29.9% | 0.95 | [0.83; 1.09] |
| **Total (95% CI)** | **556** | **755** | **692** | **751** | **100.0%** | **0.84** | **[0.64; 1.09]** |

Heterogeneity: $Tau^2 = 0.0086$; $Chi^2 = 7.55$, df = 2 (P = 0.0230); $I^2 = 72.5\%$
Test for overall effect: $t_2 = -2.88$ (P = 0.102)

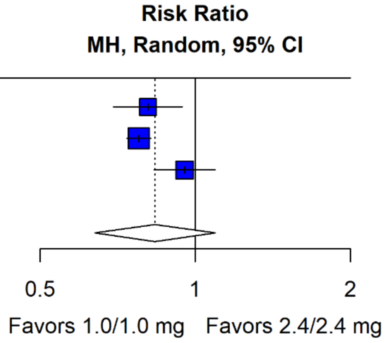


**Fig. S35** Achievement of the assigned target maintenance dose: no statistically significant difference between doses. Pooled RR 0.84 (95% CI 0.64 to 1.09; HKSJ-adjusted). Sensitivity analysis with a Wald-type CI: 0.74 to 0.95

| Studies | CagriSema 2.4/2.4 mg Events | Total | CagriSema 1.0/1.0 mg Events | Total | Weight | RR | 95% CI |
|---|---|---|---|---|---|---|---|
| Aroda 2026 | 4 | 62 | 4 | 63 | 5.2% | 1.02 | [0.27; 3.88] |
| Buse 2026 | 89 | 603 | 68 | 595 | 79.7% | 1.29 | [0.96; 1.73] |
| Rosenstock 2026 | 10 | 90 | 13 | 93 | 15.1% | 0.79 | [0.37; 1.72] |
| **Total (95% CI)** | **103** | **755** | **85** | **751** | **100.0%** | **1.19** | **[0.69; 2.03]** |

Heterogeneity: $Tau^2 = 0.0084$; $Chi^2 = 1.39$, df = 2 (P = 0.4989); $I^2 = 5.9\%$
Test for overall effect: $t_2 = 1.36$ (P = 0.307)

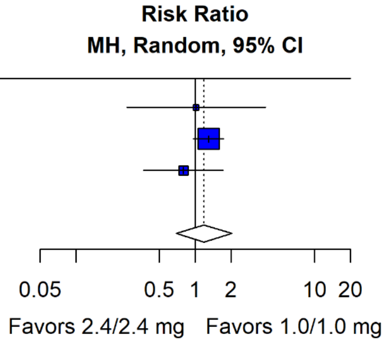


**Fig. S36** Premature treatment discontinuation for any reason: no statistically significant difference between doses. Pooled RR 1.19 (95% CI 0.69 to 2.03; HKSJ-adjusted). Sensitivity analysis with a Wald-type CI: 0.87 to 1.61
